# Automated data extraction of unstructured grey literature in health research: a mapping review of the current research literature

**DOI:** 10.1101/2023.06.29.23291656

**Authors:** Lena Schmidt, Saleh Mohamed, Nick Meader, Jaume Bacardit, Dawn Craig

## Abstract

The amount of grey literature and ‘softer’ intelligence from social media or websites is vast. Given the long lead-times of producing high-quality peer-reviewed health information this is causing a demand for new ways to provide prompt input for secondary research. To our knowledge this is the first review of automated data extraction methods or tools for health-related grey literature and soft intelligence, with a focus on (semi)automating horizon scans, health technology assessments, evidence maps, or other literature reviews.

We searched six databases to cover both health– and computer-science literature. After deduplication, 10% of the search results were screened by two reviewers, the remainder was single-screened up to an estimated 95% sensitivity; screening was stopped early after screening an additional 1000 results with no new includes. All full texts were retrieved, screened, and extracted by a single reviewer and 10% were checked in duplicate.

We included 84 papers covering automation for health-related social media, internet fora, news, patents, government agencies and charities, or trial registers. From each paper we answered three research questions: Firstly, important functionalities for users of the tool or method; secondly, information about the level of support and reliability; and thirdly, practical challenges and research gaps.

Poor availability of code, data, and usable tools leads to low transparency regarding performance and duplication of work. Financial implications, scalability, integration into downstream workflows, and meaningful evaluations should be carefully planned before starting to develop a tool, given the vast amounts of data and opportunities those tools offer to expedite research.

## Introduction

### Text Box 1: Main Contributions of this review

#### Main Contributions

- First review of automated data extraction from helth-related grey literature and soft intelligence, to automate horizon scans, HTAs, evidence maps or other secondary literature reviews.
- Includes 84 tools and methods papers mining infoirmation from helth-related news, patents, websites, trial registers, fora, or social media.
- Discusses relevant features and types of extracted data and text, level os support, evaluation, practical iomplicatios of usage, research gaps and barrieers to development and deployment of automation methods in this fild in practice.

## Background

The literature landscape in health and social care is evolving rapidly. Research outputs are being published at an unprecedented rate, which in turn has increased the rate and scale of secondary research projects, such as systematic reviews, rapid reviews, evidence gap maps, and horizon/future pipeline scans. Published and peer-reviewed literature, among other types of data, can provide important evidence used to inform choice and implementation of medicines or medical devices within a healthcare system.

However, there is a time lag between novel developments of technologies and associated research, versus their publication in peer-reviewed literature. Published research often become available years after the development of a medicine or technology. Analyses estimate the peer-review to publication time-lag for medicine and medical device trials alone as up to 4 to7 years (Blumenfeld et al., 2014; Morris et al., 2011; Van Norman, 2016); and fully relying on systematic review processes to support decision making would lead to further delays. In a recent analysis of 20,000 systematic reviews, DeYoung et al. (2021) found that the median delay between study and review publication was another additional eight years (DeYoung et al., 2021), which explains why other types of secondary literature review, such as health technology assessments (HTA) or horizon scans also use non-peer-reviewed information, to create better representations of current developments and their early evidence base (Goodman & Church, 2004; Hines et al., 2019).

To enable a comprehensive analysis of current and ongoing developments, there is a growing need to explore and consider these ‘softer’ sources of intelligence, often using a combination of grey literature and other health-related information in the public domain. Grey literature itself can be defined as “that which is produced on all levels of government, academics, business and industry in print and electronic formats, but which is not controlled by commercial publishers.”(Hopewell et al., 2007; Paez, 2017). It includes information from sources such as clinical trial registries, pre-print servers, or academic outputs such as conference proceedings, dissertations, and theses. These types of grey literature can be very valuable to health-related secondary research by uncovering first traces of new or ongoing research or collecting additional information about studies already found in peer-reviewed literature (Lefebvre et al., 2019; Paez, 2017).

However, this can also include industry-focused or legal texts such as patents or websites, or reports from governments and charities. News articles and press-releases are yet another example for public-domain and non-peer-reviewed sources of health-related information that can be considered as softer grey literature (Hopewell et al., 2007); alongside social media sites, which have the potential to provide intelligence closest to real-time development of innovations in healthcare. These latter softer intelligence sources have not been traditionally counted as grey literature.

Figure 1 seeks to illustrate the process between very early-stage research and adoption into practice on a very high level. It shows where, in theory, the scope of secondary research could be extended to include novel sources of information for an earlier detection of potentially relevant research trends. It also shows areas where automation in earlier retrieval of evidence could potentially help to accelerate discovery of information, and where automated data extraction could help to make literature review processes more efficient.

**Figure 1:**
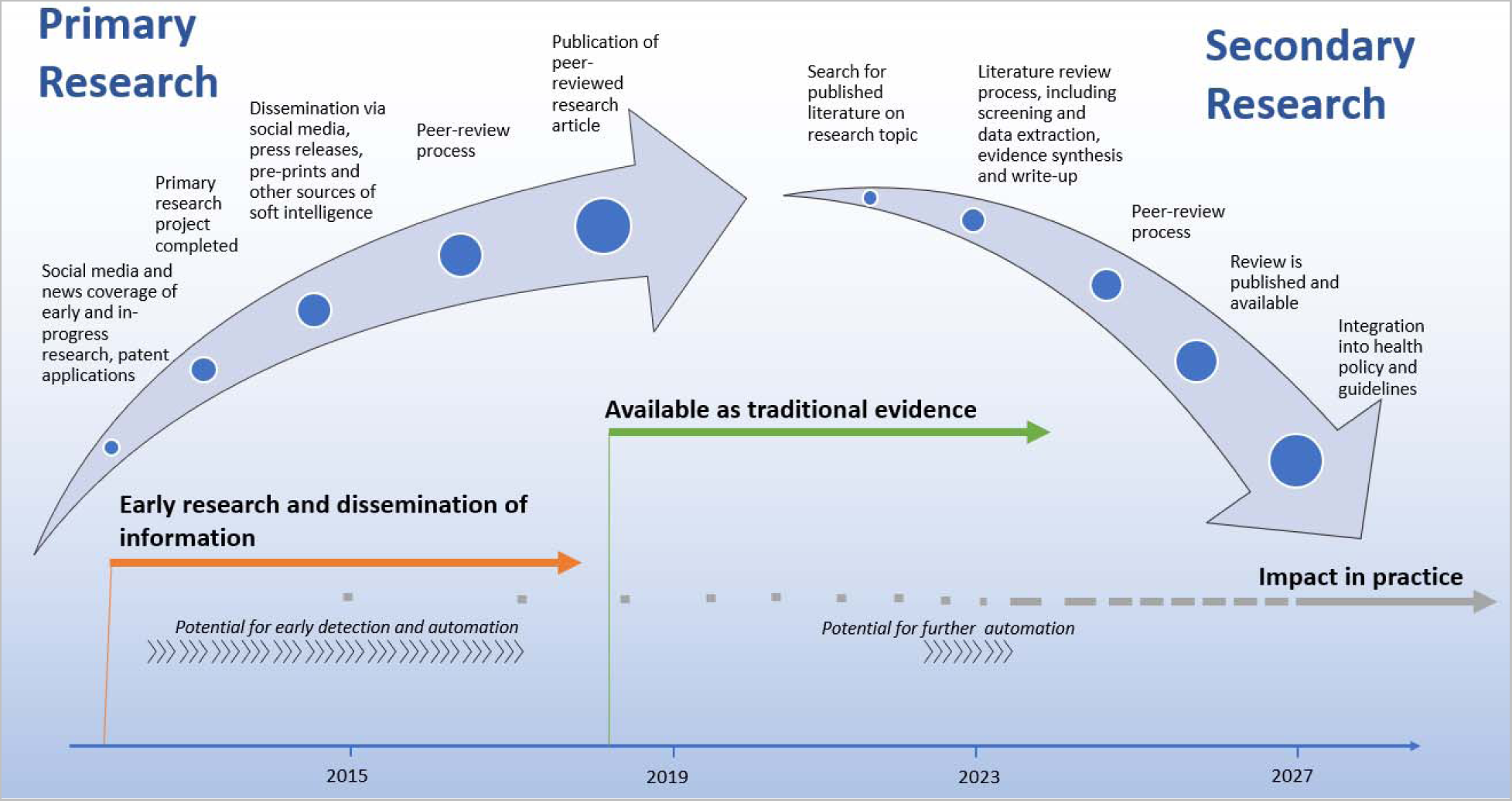
Exemplary timeline of the development and adoption of research into practice

Research areas beyond the scope of classic systematic reviews, for example horizon scanning activities or health technology assessments, utilize traditional sources of evidence (i.e. clinical trials or diagnostic accuracy studies), but also benefit from including novel sources of information from the public domain to detect signals of future trends and maturing technologies in a timely manner (Goodman & Church, 2004; Hines et al., 2019). In the scope of a systematic review, Hines et al. (2019) mapped data sources used in horizon scanning and found that softer sources of intelligence used in published projects included for example patents, surveys, or media content to detect likely technological trends in the near future.

More recently, the term ‘Infodemic’ has been used to describe the increasingly growing information landscape. The term is a neologism, combining the words ‘information’ and ‘epidemic’, thus referring to a mass of information and misinformation that is rapidly published and disseminated via social media, messaging services, or news outlets.^1^ This leads to challenges in using such data. Those challenges go back to fundamental differences between traditional, peer-reviewed evidence-based information and softer information about early research developments. The differences can be negative, in terms of lower quality of the information content (Singh et al., 2020). But they can also be positive, in terms of rapid dissemination and availability of data that would usually be held-up in lengthy clinical trials or peer-review processes.

It is critical that methods of information retrieval and data extraction advance to keep pace with this infodemic. This is especially true when expanding the scope of data sources that are used for secondary literature analysis into the domain of grey literature and soft intelligence. These information retrieval methods underpin the screening process for relevant literature and data extraction in research. Automation has a key role to play in providing faster and more resource-efficient evidence synthesis whenever the impact on general health or the implication of a medicine, therapy, or technology within a healthcare system overall is unclear.

In their 2021 survey of HTAs, which include grey literature (Goodman & Church, 2004), the WHO noted that >70% of the 127 included countries used HTAs to plan, budget, and to inform clinical practice guidelines (WHO, 2021). Stakeholders involved in the process of prioritising and creating these HTAs are government entities and national health services, as well as patient organisations or industry. However, two of the main barriers to the production of assessments such as HTAs are budget and data availability (WHO, 2021), representing bottlenecks that can be addressed via the usage of natural language processing (NLP) and automated information retrieval and extraction (Hines et al., 2019; Lauvrak et al., 2020).

## Aims

This paper provides an overview of automated data extraction methods and tools for health-related research questions that can be answered using grey and soft intelligence. We discuss the sources from which data are automatically extracted (e.g. social media, patents, news) and the type of data that are extracted (e.g. diseases, drugs, technologies). Among other items we cover performance, practical value, as well as challenges and barriers to the implementation of automated data extraction methods.

We will discuss the tools and methods within published and pre-print literature. The practical value, and robustness of these tools and the methods for automatic data extraction will be explored, based on the following three research questions:

1) What are the most important features of an extraction tool or method to support health-related research, as described in the included publications?
2) What level of support can existing published tools and methods provide to expedite evidence extraction for health-related research?
3) What are practical challenges and research gaps that constitute barriers to the development and deployment of data extraction tools and methods?

## Related research

With advances in NLP and developments in deep– and machine-learning it is becoming feasible to process vast amounts of unstructured digitalized texts. This is giving rise to the emerging field of NLP-based health data science, where novel research in data mining and data extraction is specifically applied to automate work in evidence synthesis. A living systematic review of automated data extraction from the highly related field of peer-reviewed health literature currently includes 76 papers, indicating fast-paced advancements in the areas of automatic extraction, normalisation, relation extraction and text summarisation (Schmidt et al., 2021). Within these advancements there remains a need to explore methods of automatically processing unstructured text data in the non-peer-reviewed space, and to assess which tools and methods will facilitate this process for end-users. NLP and text mining is frequently used to analyse or extract data from social media platforms such as Twitter (Acosta-Urigüen et al., 2020). Applications range from vehicle traffic analysis (Acosta-Urigüen et al., 2020) to medical and health data (Viviani & Pasi, 2017). Correia et al. (2020) published a narrative review of recent work on data mining in social media content analysis. They discuss papers on automation in the domains of pharmacovigilance and sentiment analysis, most commonly targeting specific drugs and their adverse events, or mental health research questions. A large amount of related research has been conducted on information extraction from electronic health records, for example extracting diagnoses, treatments (Xiao et al., 2018), or genomic data (Miller & Shalhout, 2021). Other grey literature data sources such as pre-prints (Cabanac et al., 2021) and clinical trial registrations are targets for data mining and extraction to connect them with their published counterparts (Liu et al., 2022; Smalheiser & Holt, 2022).

## Methodology

### Research objective

This review maps published tools and methods for literature mining and data extraction. A ‘tool’, in this context is defined as an end-user application with a user-interface, available for example as web– or desktop application. A ‘method’ is defined as a set of scripts or a description of an algorithm that requires users to be familiar with data science or programming. Results of this literature review were summarised in the form of an evidence map, visualizing the extracted data, current knowledge, and research gaps. The review includes any publications that describe approaches to expedite data extraction from grey literature and soft intelligence. For the purpose of this research, grey literature and soft intelligence includes any health-related data that has not passed peer-review; with examples given in Table 1.

**Table 1:** Examples for types of grey literature information and exemplary data sources

| Information type | Examples for data sources |
| --- | --- |
| Social media | Twitter, Reddit, YouTube |
| Internet fora | Mumsnet, SANE |
| News | Google News, Med-Tech News, PharmaTimes |
| Government Agencies or charities | Websites, eg UNICEF, world bank |
| Patents | Databases and indexes, such as USP |
| Clinical trial registries | ClinicalTrials.gov and other registries |
| Pre-prints | MedRxiv, ArXiv |

**Table 2:** End-user tools for automated data extraction. All references are tagged and available within the SWIFT-Review project file under the “Tools” tag.

| Title | Tool name | Data | Deployment | Reference | Source | Link to tool |
| --- | --- | --- | --- | --- | --- | --- |
| **A user-friendly tool for medical-related patent retrieval** | TWINC | Patents | Unclear | Pasche, E., Gobeill, J., Teodoro, D. et al. | PubMed |  |
| **PADI-web 3.0: A new framework for extracting and disseminating fine-grained information from the news for animal disease surveillance** | PADI-web | News | Web | Valentin, S., Arsevska, E., Rabatel, J. et al. | PubMed | https://padi-web.cirad.fr/en/ |
| **A new visual navigation system for exploring biomedical Open Educational Resource (OER) videos** | - | Video | Unclear | Zhao, B., Xu, S., Lin, S. et al. | PubMed |  |
| **Mining Adverse Drug Reactions from Unstructured Mediums at Scale** | - | Social Media, EHR | Unclear, some code given | Ul Haq, H., Kocaman, V., Talby, D. | ArXiv |  |
| **Development and evaluation of a prototype search engine to meet public health information needs** | PHIS | Websites | Unclear | Keeling, J. W., Turner, A. M., Allen, E. E. et al. | PubMed |  |
| **iPresage: An innovative patent landscaping tool** | iPresage | Patents | Web | Avasarala, V., Bonissone, P. | Scopus |  |
| **E-patent examiner: Two-steps approach for patents prior-art retrieval** | E-patent examiner | Patents | Web | Kravets, A. G., Korobkin, D. M., Dykov, M. A. | Scopus |  |

**Table 3:** Level of automation in the included papers. All references are tagged and available within the SWIFT-Review project file when creating a publication year heatmap with the “Extend” tag. One paper can include more than one type of automation.

|  | 2005-2006 | 2007-2008 | 2009-2010 | 2011-2012 | 2013-2014 | 2015-2016 | 2017-2018 | 2019-2020 | 2021-2022 |
| --- | --- | --- | --- | --- | --- | --- | --- | --- | --- |
| Priorisation and Summarisation | 1 | 0 | 2 | 5 | 3 | 8 | 15 | 16 | 8 |
| Mining Entities and Sentences | 0 | 0 | 0 | 1 | 3 | 10 | 8 | 15 | 8 |
| Extraction and Normalisation | 0 | 0 | 0 | 0 | 1 | 2 | 6 | 5 | 3 |

Considering open questions around usefulness, feasibility, and practical integration of grey literature and soft intelligence, the motivation for this literature review is to identify and examine tools and methods that currently exist and have been used to automate data extraction activities from these publicly available data sources.

### Literature searches

A robust search strategy was developed to identify relevant articles from a variety of electronic databases, covering health, informatics, and pre-prints in both health research and informatics. The initial search strategy was developed using the PubMed ‘Advanced Search’ function. Six databases were searched (2005-2022), each using a database-specific adaptation of the PubMed search: MEDLINE (searched via PubMed); Scopus; ACL; dblp computer science bibliography; MedRxiv; and ArXiv.

| Database | Interface or URL |
| --- | --- |
| MEDLINE via PubMed | https://pubmed.ncbi.nlm.nih.gov |
| Scopus | https://www.scopus.com |
| ACL Anthology | https://aclanthology.org/ |
| dblp: computer science bibliography | https://dblp.org/ |
| MedRxiv | https://www.medrxiv.org/ |
| ArXiv (computer science) | https://arxiv.org/archive/cs |

The start date of 2005 was selected, since this is the year after which publications relevant to text-mining in general systematic review automation first started to appear. Three published systematic reviews of data extraction methods from the related field of peer-reviewed literature did not find any published text-mining or data extraction approaches prior to 2005 (Jonnalagadda et al., 2015; O’Mara-Eves et al., 2015; Schmidt et al., 2021). We furthermore decided to keep this 2005 date filter because the availability of data sources changes over the years, and methods published prior to this date are not representative anymore or are becoming unlikely to be usable in practice due to changes and updates in programming languages (ie. new Python^2^ or Java^3^ versions).

The PubMed search strategy was developed, and refined further based on feedback from an independent information specialist. The strategy was then adapted for usage in Scopus. Searches on the ACL, dblp, MedRxiv and ArXiv were adapted and carried out as described by McGuinness and Schmidt (2020). In short, we utilized full database exports of all papers indexed by these databases, and then used methods from the medrxivr R package^4^ to retrieve relevant records. Search strategies, including the regular-expression-based search for the ACL and pre-print servers, are attached as Appendix D at the end of this document.

### Eligibility criteria

On the title/abstract and full text level, we included papers that met the following criteria:

1. Original publication about the development of an original data extraction tool or method.
2. Data subject to automatic processing include non-peer-reviewed data related to healthcare. On full text level, we added further inclusion criteria:
3. Papers available as full-text publications. We included journal articles, conference papers, or pre-prints.
4. Publication in English.

We excluded papers that met the following criteria:

1. Tools and methods that extract data from patient’s electronic health records, discharge summaries, death certificates or any type of individual patient level data.
2. Tools and methods that extract genomic or biological data such as gene expressions or proteins.
3. Tools and methods that are developed or evaluated with a focus on data from peer-reviewed texts only, in the absence of using any grey literature data.

In addition, to these inclusion criteria, during title and abstract screening we separately tagged papers that describe the usage or evaluation of a tool or method with respect to a specific health research question. For this, we tagged two items:

1. The topic of research: We created a vocabulary to categorize and bin the specific health topic studied in the reference, based on information available in the title and abstract. The tags included, for example, mental health or Covid-19.
2. The data sources: We created a vocabulary to categorize and bin the sources of mined data. The tags included, for example, Twitter or health-related fora.

The decision to tag, but then exclude topic-specific research papers at the title/abstract level was made after a pilot-study showed that full inclusion of every such paper would lead to an unfeasibly large amount of included papers. We imported all tags into the SWIFT-Review software (Howard et al., 2016), to create visualizations in the form of heatmaps, bar– and pie-charts and to make the whole dataset publicly sharable. A description of these results is given in Appendix C.

### Screening and workflow management

All papers were deduplicated, screened, and data-extracted in SWIFT-ActiveScreener (Howard et al., 2020). Screening at title/abstract level was conducted up to an estimated sensitivity of 95% by one reviewer, and a second reviewer independently checked random samples of in– and excluded records. All conflicts were discussed and resolved until the screeners were confident that the in– and exclusion criteria were applied correctly.

Similar to the initial screening process, full-text screening and data extraction decisions were reviewed by an independent reviewer. Conflicts were discussed and resolved in the same fashion.

Where there were multiple publications describing the development or evaluation of the same tool, we grouped those papers and jointly extracted data once for each tool, focusing on the most recent version of each feature or function.

## Data extraction

Data were extracted within SWIFT-ActiveScreener. For each research question (RQ), the data extraction questionnaire was set up in the form of text fields and checkboxes, as applicable. In the following we provide an overview of the extracted data, the full questions are shown in Appendix A.

**RQ1: What are the most important features of an extraction tool or method to support health-related research?**

We extracted relevant features and functionalities from each paper, a list of data sources from which information was obtained (eg. GoogleNews or Twitter), and the type of data (eg. patents or online fora)

**RQ2: What level of support can existing tools or methods provide to expedite evidence extraction in health-related research?**

We extracted whether the paper refers to a tool or method, the extent of automation (e.g. recognition of entities or full normalisation to standardised vocabularies), metrics and methods used for validation within the paper (e.g. F1, precision), and description of the tool’s integration into real research projects, where applicable.

**RQ3: What are practical challenges and research gaps that constitute barriers to the development and deployment of data extraction tools and methods?**

We extracted any challenges or barriers related to the development or deployment of tools and methods, caveats when using these in real-world research projects, and research gaps described in the papers.

## Results

### Screening

| Database | Number of results |
| --- | --- |
| PubMed | 2,108 |
| MedRxiv | 89 |
| dblp | 81 |
| ArXiv | 1,332 |
| ACL | 387 |
| Scopus | 5,704 |

In total, the searches retrieved 9,701 references (databases searched up to 03/2022). After deduplication 8,927 references were imported into SWIFT-ActiveScreener for screening.

As described within the method section of this paper, screening was conducted using early stopping at a target estimated sensitivity of 95% (Howard et al., 2020). After reaching the target sensitivity a further 1000 references were screened, but screening was then stopped because no further relevant records were identified.

In total 3646 titles and abstracts were screened. Of those, 318 titles and abstracts were excluded at title and abstract level, but still tagged by research topic and data source because they described topic-specific analyses conducted based on non-peer-reviewed data. These tags were only applied during the abstract screening process, and the 318 references did not proceed to full text screening.

On full text we included 83 tools and methods. 84 papers were included, but two were grouped together because they described the same tool.

### Summary of the full-text literature

A total of 84 papers for 7 tools and 76 methods were included at the full-text level. One tool was described by two papers. Data to answer the research questions was extracted into extraction forms created within the screening tool.

#### RQ1: What are the most important features of an extraction tool or method to support health-related research?

A total of 16 papers described features and functionalities implemented in tools, or features that could be of use in practice when processing the data via a method. Pasche et al. (2012) noted the ability to bulk-process data and the utility of automatic query expansion, for example to automatically increase the amount of chemical terms by adding synonyms found within MeSH or Pubchem terminologies within their tool called ‘TWINC’. As part of the papers for the PADI-web tool, Valentin et al. (2021) described the feature of automatic daily evidence updates to the data, via RSS feeds, crawling of related websites, and usage of the Google News API. The tool can retrieve new data and therefore prioritise, mine, and normalise new information as it is published; ensuring that research-projects are up-to-date. They support data annotation via an integration of the BRAT tool (Stenetorp et al., 2012) and make the tool publicly available as web-application, thus facilitating collaboration and re-use of manually extracted data. They furthermore describe features such as email-notifications and summaries sent to the user, which helps with transparently communicating changes in the evidence and providing fast and easy-to-digest updates without accessing the tool itself every day. Hariprasad et al. (2015) discuss added value of a user-interface to visualize automatically mined or extracted data, in the form of histograms, pie charts or other types of plots.

Natsiavas et al. (2019) describe the user-requirements and design-process of a future tool, citing the full pipeline of prioritisation/mining/normalization as a feature. They discuss the problem of heterogeneity between data sources and suggest a division to explore data from different sources separately, as well as separate data mining and normalization for each data source. As final, separate feature, they describe a data consolidation process that includes automated reports and visualisations to follow-up on new data. In the PHIS tool, which stands for Public Health Information Search (Keeling et al., 2011), public-health websites are crawled and there can be a focus on more than one class of entities. Documents summaries are provided with respect to extracted data. Tafti et al. (2017) describe a data mining architecture for mining social media data, and note that usage of their database-infrastructure as a feature to increase scalability and future access via a tool.

Lee and Uzuner (2020) processed patent data and described benefits of a feature to divide patents between already-commercialized products and between technologies in development. Also in the patent-space, Avasarala and Bonissone (2012) (iPresage tool) describe colour-coding patent-assignees for better identification and visualizing temporal trends via stacked histograms. The E-patent examiner tool by Kravets et al. (2016)is also a patent-focused web-application, citing being web-based as a positive feature, as well as allowing expert-input on top of the automated process.

For video-data, Zhao et al. (2016) (unnamed tool) describe implementing features that help users gain a streamlined overview of the data. This includes automatically indexing and updating their dataset with new health-related videos, similar to the updates within PADI-web. They also mention benefits of making the tool available as web-application. The video-specific features include visualizing mined content as part of the video timeline and making it easy to skip between highly relevant sections, using hover text and visual cues such as word-clouds that represent key moments. Multiple videos can be visualized on the same grid for comparison. In terms of user-management, they discuss features to add comments to a video and user-account management.

### From which source are data retrieved and extracted?

We tagged the source of data used within the included full-texts and show the results in Figure 3, using the same set of tagging categories that was also applied to the topic-specific datasets analysed on title and abstract-level in Appendix C. The results within Error! Reference source not found. (topic-specific abstracts) and Figure 3 (included full-texts) are very similar, both indicating that Twitter, health-related fora, websites and news are the most common sources of data used for automation.

**Figure 2:**
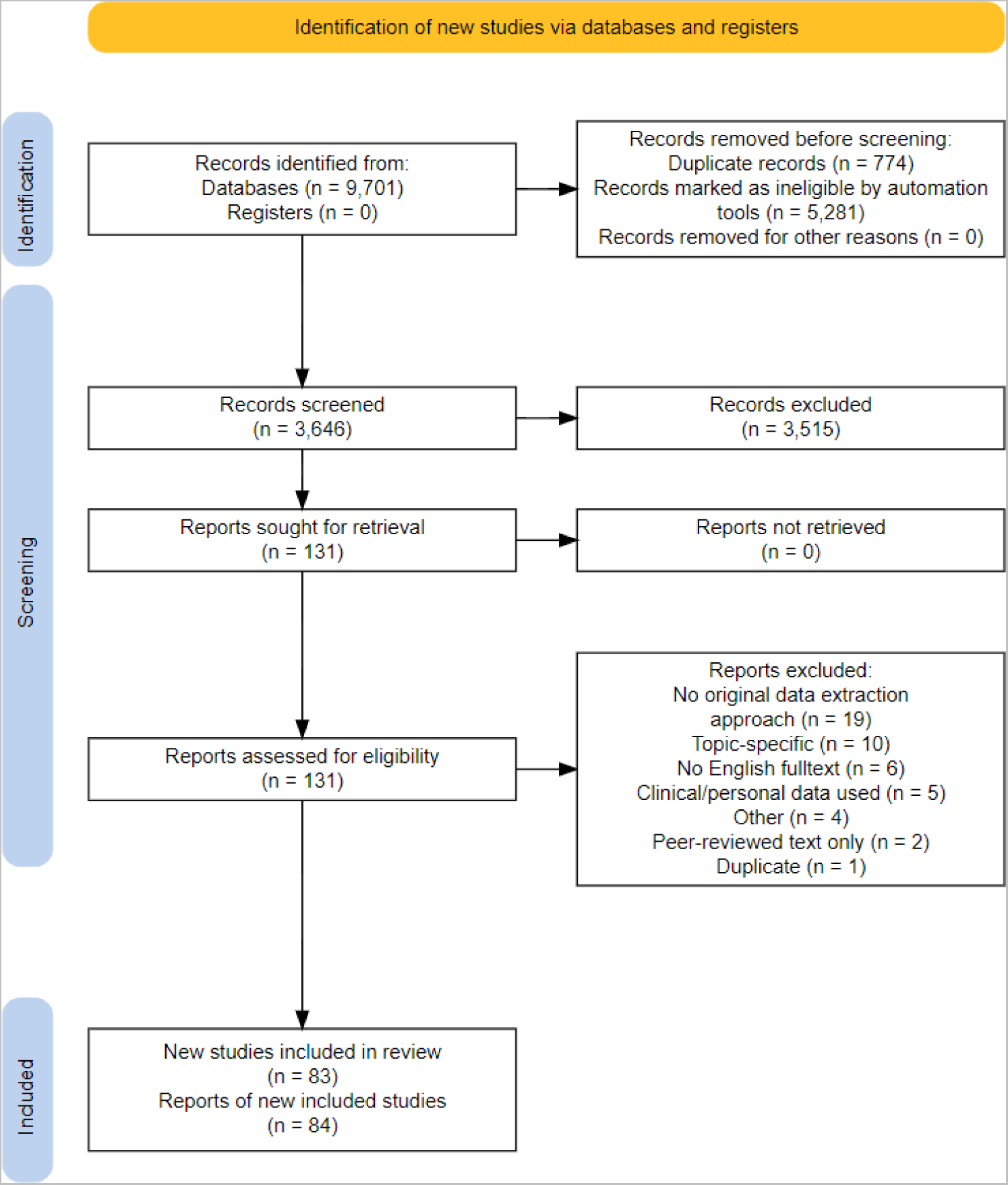
PRISMA2020 flow diagram (Haddaway et al., 2022; Page et al., 2021)

**Figure 3:**
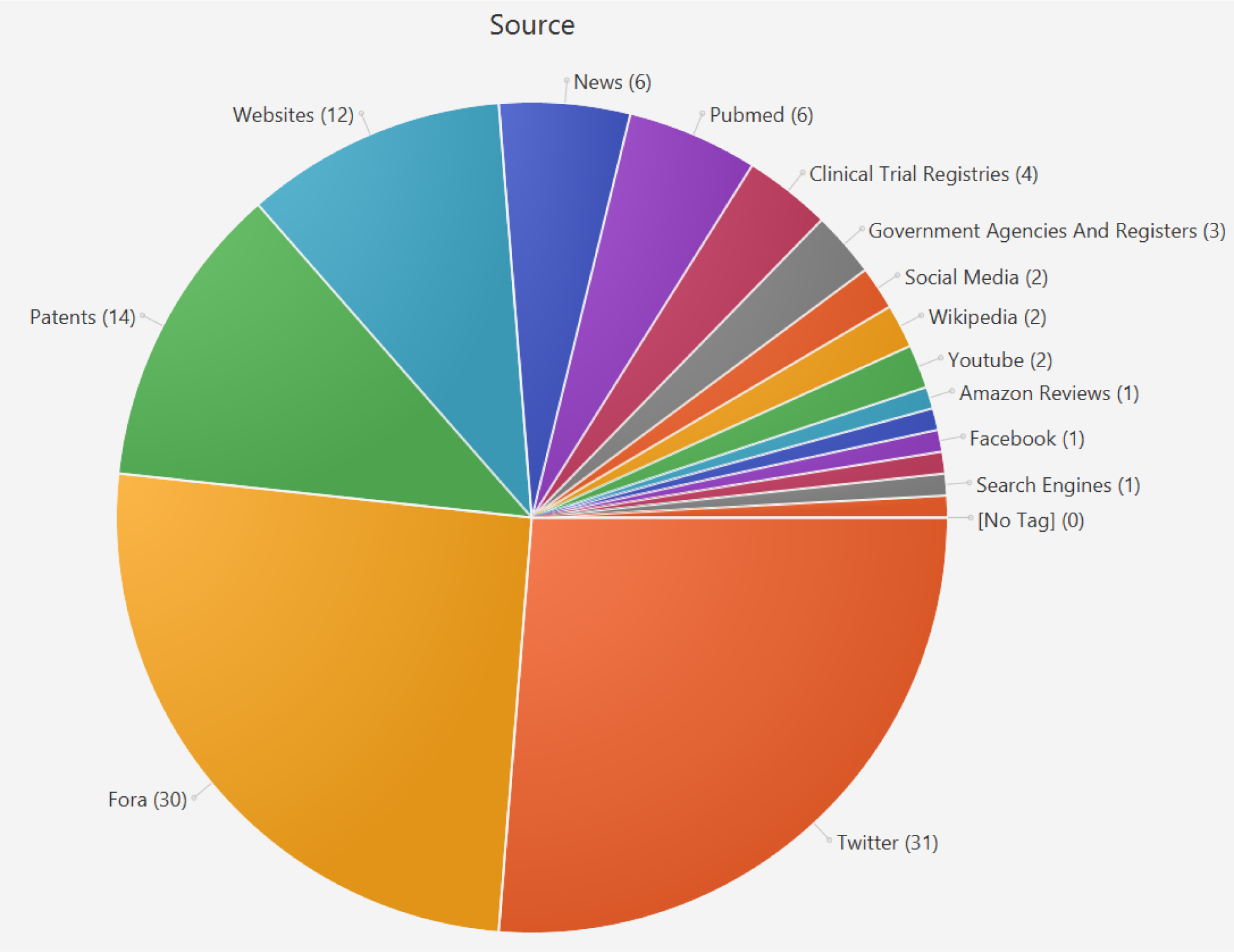
Sources of data used in the included papers. We only included full-texts if they reported usage of such data within their dataset. PubMed was tagged as data source whenever an included reference mentioned using a mixed corpus such as TwiMED, including a mix of PubMed and Twitter data. References within each category can be accessed within the SWIFT-Review project under the “Source” tag. One paper might include more than one source of data.

For the purpose of training and evaluating an algorithm, researchers often use publicly available benchmarking corpora or they create custom labelled datasets for this purpose.

We found 25 different benchmark corpora used as data sources for training and/or evaluation in the included papers. The most commonly used corpora were SMM4H^5^ used by 8 papers, and CADEC (Karimi et al., 2015) and TwiMED (Alvaro et al., 2017), used by 5 papers each. One corpus was used by two papers, and the remaining 21 corpora were used by only one reference each (see Figure 4).

**Figure 4:**
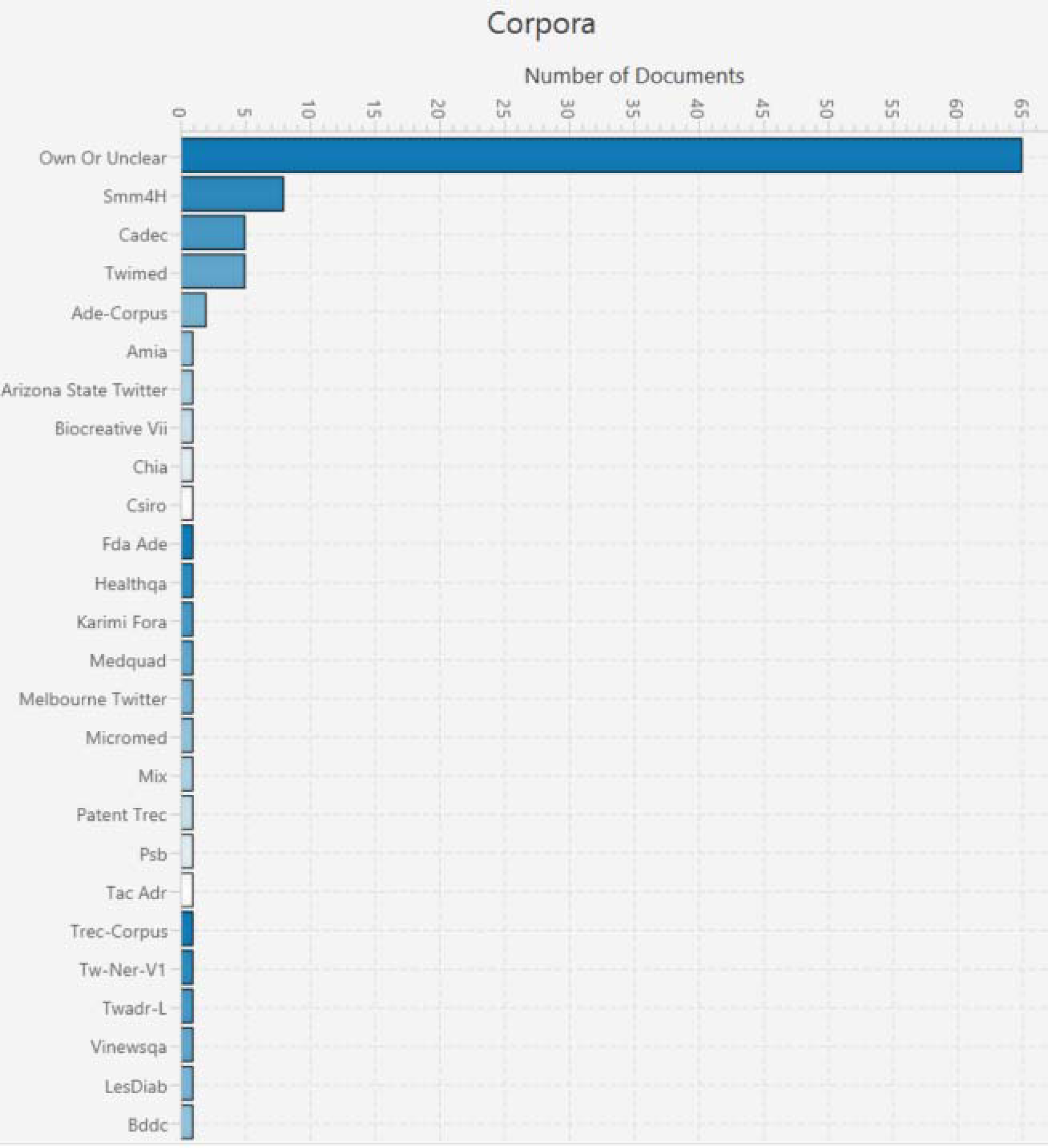
Corpora used in the included tools and methods. All papers with their corpora can be filtered within the SWIFT-Review project, under the tag ‘Corpora’. One paper might include more than one corpus.

From the 84 papers, at least 65 reported creating their own datasets. This included either curating a full dataset from scratch or labelling and using a smaller dataset in addition to a previously published benchmarking dataset. The high usage of own and custom datasets can in part be explained by the heterogeneous characteristics of non-peer-revied data.

Peer-reviewed literature itself is commonly published in English. In contrast to that, information extracted from publicly available and grey literature sources is much more diverse. Social media posts, forum discussions or data from government agencies are often available in native languages of the authors who conduct the NLP research. We found examples for Indonesian (Halim et al., 2018), Chinese (Chen et al., 2019; Zhao et al., 2017), Croatian (Kocijan et al., 2020), and other multilingual datasets including French or Latin (Grabar & Hamon, 2014). This diversity in both data sources and in languages leads to a greater need to curate datasets on a project-by-project basis.

Currently, the biggest publicly available datasets include tweets and forum posts mostly in English, while availability of multilingual data is more limited. Datasets are often small and include imbalances within the labelled classes, which in turn makes it difficult to train reliable and well-performing algorithms for automation(Saha et al., 2020). Saha et al. (2020) also suggest that future research into using well-trained and evaluated methods for automatic translation into English is warranted. The performance of machine-translation and so-called multilingual zero-shot classification has improved greatly in recent years, but its application to data extraction for medical contexts has not been evaluated in great detail due to a lack of multilingual corpora.

### Which types of data are extracted?

Adverse events (n=35), followed by disease (n=29), and drugs (n=18) were the most common types of data addressed by the automation models in the included tools and papers (see Figure 5). We tagged a total of 32 different types of data that are commonly used within health-related literature analyses. Some of these, such as the 14 papers categorised under ‘Technology and Trends’ can be of use to researchers implementing horizon scanning automation, while others such as ‘Symptom’ (n=16) or ‘Intervention’ (n=14) may be useful for a variety of literature review questions and methodologies.

**Figure 5:**
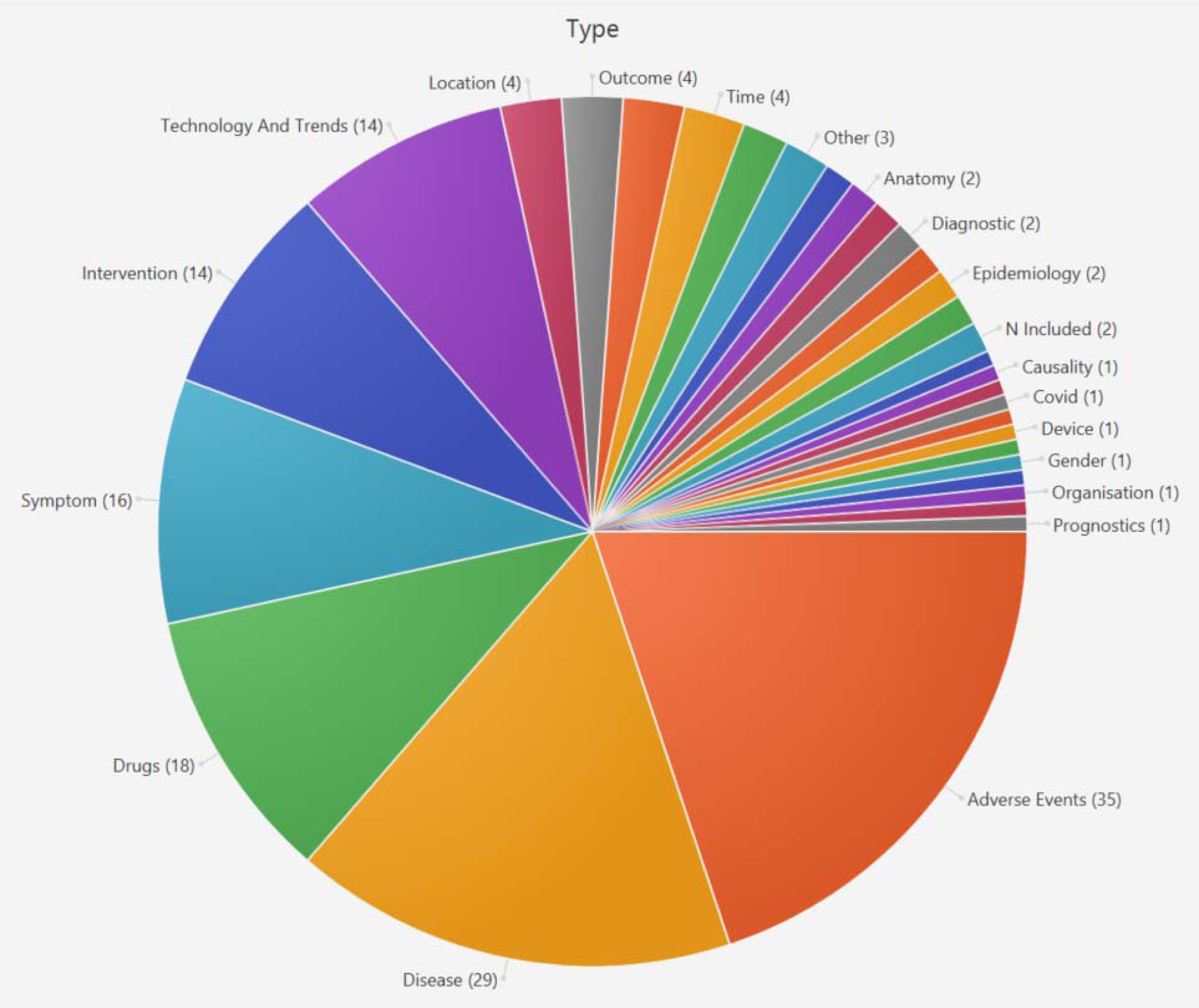
Types of data covered by automation methods in the included tools and papers. All papers can be filtered and accessed via the SWIFT-Review project under the “Type” tag.

#### RQ2: What level of support can tools and methods provide to expedite data extraction in health-related research?

##### Does the reference describe an end-user tool?

We found 7 end-user tools and 76 published methods papers. For one tool (PADI-web) we aggregated two full texts into one tool description (Rabatel et al., 2019; Valentin et al., 2021). Those tools automate data extraction across different types of text (i.e. patents, news, trial registrations) and across different media (i.e. digitalised text and videos). For PADI-web we found accessible web-deployments, giving users the opportunity to test and use the tool. Code has been published for one further tool’s NLP models. The remaining tools were not accessible online and we were unable to find publicly accessible deployments or executable desktop applications.

The table below shows an overview of the tools, key information about them and a short description.

### To what extent is data extraction automated?

For each included paper, we tagged the extent of automation based on three options. These are explained in more detail below, and key aspects are further described in the glossary, which is given in Appendix B.

– Prioritisation and summarisation of evidence (lowest level of automation)
– Mining entities and sentences
– Extraction and normalisation to standardised vocabularies (highest level of automation)

### Prioritisation and summarisation of evidence

In the research context of this review, priorisation and summarization is very similar to the well-known NLP task of document-level classification and topic modelling. We found 63 papers that described the functionality of helping researchers to summarise, re-order or identify whole documents related to a health-related research question or task. This can be achieved by classifying whole tweets by content type or by identifying emerging technology trends within YouTube video captions or patents. For example, a tweet could be classified and prioritized as containing content about adverse events in general. This process helps to streamline the identification of relevant content by presenting researchers with a pre-filtered set of likely-relevant research. Documents, as such, are not being data-extracted but rather pre-sorted and prepared for analysis. This process was included in the scope of this review paper because it is generally regarded as one of the most straightforward applications of AI and machine-leaning, with a chance for high, reliable model performances. In the domain of screening peer-reviewed papers for systematic reviews, using AI in the prioritisation process is now widely recognized approach to save significant amounts of time in order to find relevant papers (Howard et al., 2020; O’Mara-Eves et al., 2015).

Out of the 63 papers, 25 added more value by combining the prioritisation step with the more specific task of mining entities or sentences, and 13 of those papers also covered the whole process of creating structured data by adding normalization functionalities. In the following paragraphs, we describe the tasks of mining and normalization in more detail, to give an overview of those processes and their potential added value. As part of this evidence map, we share a SWIFT-Review project that contains all included papers and the tags discussed in this section, such that readers can browse the papers in each category easily.

### Mining entities and sentences

Here we tagged papers if they described processes that lead to the targeted identification of shorter pieces of information in text, for example sentences, named entities, or relations between them. An entity could be a single word or short phrases of text belonging to a clearly defined class of things, such as the word ‘Aspirin’ being an entity of the class ‘Drug’.

Tasks related to data mining are harder than the prioritisation or summarisation, because they often require classification on a word-by-word basis and thus introduces a higher chance of errors or partly-correct identification of entities. The input text was usually natural language, in full texts or segmented into units such as sentences, abstracts, or paragraphs. In the included papers, 46 reported some form of mining functionalities within their text, mostly limited to named-entity recognition and not focussing on relation extraction. In practice, this leaves the user with selected pieces of text in a semi-structured form, because the resulting text is shorter and has class-assignments, such as ‘drug’ or ‘disease’. However, the mined text itself is just a subset of the original text, and therefore still present in the form of natural language. This natural language can carry variations in expressions that complicate automated synthesis of the data, thus still requiring human assistance and downstream manual work.

### Extraction and normalisation

To create fully structured data from unstructured text, all mined text can be normalised to a structured vocabulary. For example, when normalising to MeSH terminology, mined text pieces such as ‘2-(Acetyloxy)benzoic Acid’, ‘Polopiryna’, or ‘acetylsalicylic acid’ would all be resolved to MeSH term ‘D001241: Aspirin’. In total, 17 papers described normalisation as part of their pipeline. The task of normalisation is harder than mining entities or sentences, because the core-classification task is not binary (ie. not a choice between the decision drug/not-drug for a word) but rather a complex multi-class and sometimes multi-label case where the potential decision-space is as large as the vocabulary to normalise to. For example, when normalizing to MeSH terms, there are more than 680,000 entry terms that can be chosen to normalise an entity to. This does not only create computational problems because of the large space of potential labels, but also problems in terms of ambiguity, non-covered vocabulary, and variations in specificity of the chosen concepts. In other words, one mined piece of text may correctly refer to one, more than one, or to no covered concept within a vocabulary. Whenever more than one correct concept applies, one might need to make the choice between less specific normalisations (ie. high-level concepts in the MeSH tree) or more specific normalisations (ie. the lowest-level finer-grained concepts). This increases not only the complexity of the classification task, but also makes it challenging to conduct a fair and comprehensive evaluation that is representative of future, unseen data that will be seen by the system when it is deployed in practice. Furthermore, in practice, a correct normalisation requires correct named-entity recognition in the first place, thus escalating any errors made during downstream data processing. This accumulation of error during multiple classification-steps is a challenge that may be further reducing the amount of correctly normalised entities when tools and methods are used in practice.

### Metrics and strategies used for the evaluation of algorithms

Evaluations were most commonly performed in a quantitative manner by using manually or distantly-labelled gold-standard datasets. Most commonly the process included the creation of a dataset by experts, and then splitting data randomly into training, validation, and test sets, to ensure that none of the data seen by an algorithm during the training-phase is used for evaluation. The process of splitting the data was either described in the papers, or authors described using published benchmark-datasets with predetermined splits. In line with scores frequently used to report automated data extraction results on peer-reviewed literature (Schmidt et al., 2021), the commonly used evaluation metrics of precision, recall, and F1 score were the most prevalent scores, with 19 papers reporting all three scores. F1 by itself was the most common score, reported in 47 papers, followed by precision (n=29) and recall (n=26). Accuracy was reported in n=9 papers, MAP in n=4, area-under-curve n=3, specificity n=2, one devised a new score and two papers used speed (see Figure 6).

**Figure 6:**
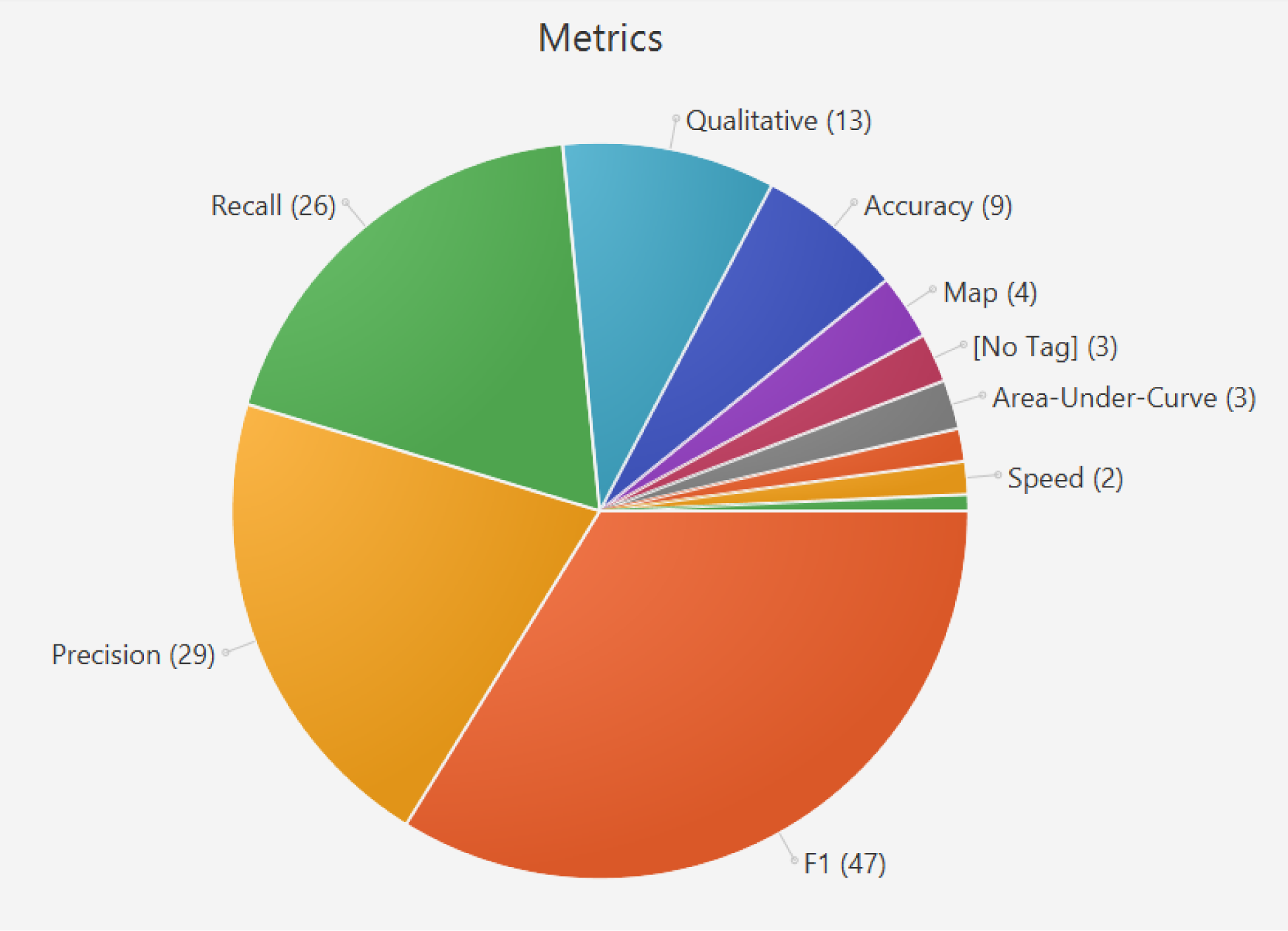
Level of automation in the included papers. All references are tagged and available within the SWIFT-Review project file under the “Metrics” tag. One paper can include more than one metric for evaluation.

Those scores were reported across different kinds of classification tasks, generally showing very good scores for straightforward tasks that include only document-level classification. For these binary classification tasks F1, precision, and recall scores higher than 0.9 is becoming more common (Atal et al., 2016; Ellendorff et al., 2018; Fan et al., 2020; Guo et al., 2017; Rezaei et al., 2020). Scores decrease for harder classification tasks such as normalization to controlled vocabularies, commonly ranging between 0.2-0.6 in precision, recall or F1. In part, this large variation between reported scores, spanning 0.2-0.6, can be seen for normalization because the included papers used the same evaluation metrics but applied them in different ways. An example for this is the usage of relaxations such as counting a predicted answer as ‘correct’ if the true label was predicted within the top-N predictions, as opposed to only accepting one answer. Another relaxation method was to decrease the amount of potential labels to only include top-level categories (Grabar & Hamon, 2014; Magge, O’ Connor, et al., 2021; Saha et al., 2020; Zhao et al., 2017).

Even when grouping and comparing classification scores for the three classification tasks separately it is not straightforward to determine the best-performing algorithm within each category. Algorithm performance reported on a domain-specific dataset labelled by a group of researchers according to their own annotation guidelines may vary when the same algorithm is tested on completely new and therefore independent data labelled by different persons. A number of included papers used more than one dataset to evaluate their classifiers in parallel, showing differences between the evaluations scores of the same algorithm or architecture and therefore making it hard to estimate how each algorithm would perform in real-life, with potentially new or evolving data (Arnold et al., 2020; Guo et al., 2022; Korkontzelos et al., 2016).

In total 15 papers described qualitative or practical evaluations of their algorithms. In the most cases, the qualitative evaluation completely replaced quantitative analysis. Evaluations were conducted via case-studies and explorative analyses of large sets of unlabelled data. By applying the algorithm or proposed tools to a real-life dataset, authors discussed their perceived value of the automatically mined or extracted data. In the field of mining emerging health technologies, for example, it was a common approach to train Latent Dirichlet Distribution (LDA) topic model algorithms to identify dominant themes in large corpora of data (Daniel & Dutta, 2018; Guo et al., 2017; Sofean & Aras, 2018; Zhao et al., 2017; Zhou et al., 2021). This approach is chosen because LDA is a generative unsupervised machine-learning approach, assigning a pre-defined number of topics to each document and using probability distributions across the vocabulary to assign words to topics (Blei et al., 2003). The output of this algorithm is a set of unlabelled word-clouds with vocabulary that may have emergent semantic similarities when examined by a human. Those outputs are then qualitatively evaluated by picking emerging topics and discussing or visualizing data. Drawbacks for using topic-models such as LDA are that they require human interpretation, they are strongly influenced by training parameters (such as the pre-defined number of topics per document) and in the absence of a fully labelled dataset it is not possible to estimate sensitivity (i.e. to estimate the amount of missed important topics). However, barriers to conducting a full quantitative evaluation can include, for example, a lack of resources to create own labelled datasets where none are publicly available. For this reason, researchers are opting for such unsupervised algorithms to create value, without investing large amounts of resources (Rastegar-Mojarad et al., 2016).

Other practical evaluations in the absence of labelled data include assessments of efficiency within the workflow, by doing a direct comparison between time-taken by humans to complete a task versus AI-supported humans receiving automatically extracted or likely relevant data first. Zhao et al. (2016), for example, found that using their unnamed AI tool in explorative analysis of biomedical video content increased the speed of finding relevant evidence by 3.7 times, and that users were able to answer content-based questions more accurately. Keeling et al. (2011) qualitatively compared their search tool PHIS with Google, applying the ‘Critical Incident Technique’ by asking users to keep notes of situations where the tools were effective or ineffective, and conducting retrospective semi-structured interviews.

### Description of an automation method’s integration into current review workflow, as described within each paper

Four included papers described using their tool or method in practical settings. Tasks in which a tool or method was used included information gathering and scoping before starting a review project, usage during the data extraction phase, or in the scope of clinical practice.

The PADI-web 3.0 tool (Valentin et al., 2021) describes integration into practice both in terms of scoping and in terms of keeping researchers up-to-date automatically, on a daily basis. When describing the scoping process, they note that one way to integrate automation of grey literature data extraction is by using it to ‘triage’ information before further review. Triage itself is a term describing prioritisation of patients in emergency situations, to administer treatment first to anyone who might benefit from it most. Similarly, researchers in health-related topics focus on the best available, peer-reviewed evidence first. But then, a system of triage for less strong sources of evidence, such as news and social media, may be applied. Especially in the field of disease surveillance, Valentin (2021) note that classifying and removing likely irrelevant data from additional sources of information via machine-learning is a step that brings value to a research project because it leads to a prioritized and therefore earlier detection of information. The practical integration into review workflows happens via automated dissemination of content to the reviewers. Specifically, automated email-updates, summarization of newly classified relevant information, and usage of RSS-feeds is described (Valentin 2021).

Similar to PADI-web 3.0, Zhao et al. (2016) describe a tool for information scoping that is not directly connected to feed information into review tools. Using biomedical educational videos, they support the process of scoping by helping to apply information-specialist curated keyword searches and skimming video content. In practice, this aims to prioritise the display of likely relevant content and thus reducing time needed to watch the full content. However, similar to PADI-web, no export of the classified or extracted information into downstream evidence-synthesis tools is described.

Turner et al. (2005) developed a data model describing key pieces of evidence extracted from grey public-health data, for downstream usage in automation tools. They characterized information needed in practice and discuss using a rule-based approach for automatic information extraction that firs their data model.

In contrast to the common focus on literature reviews, Natsiavas et al. (2019) discuss requirements for the integration of new information into a clinician’s workflow. They do not describe integrating with downstream tools, but describe value added for clinicians by providing automated summaries and analyses of texts describing adverse events. They mine data, normalise and consolidate, provide structured reports and follow up on new data extracted from social media, government websites such as FDA, and patient health records.

#### RQ3: What are practical challenges and research gaps that constitute barriers to the development and deployment of data extraction tools and methods?

##### Challenges or barriers that hinder the development or deployment of new tools, as described in each paper

###### Heterogeneity and transferability within the data

Heterogeneity within the data was discussed as a barrier to the overall process by Sofean and Aras (2018). They focused on patent mining and noted that within patent documents there are different kinds of text such as the main text and metadata, including information about the inventors, institutions, and timelines. The challenge of automating data extraction from patents exceeds the boundaries of NLP, because they include potentially valuable information in forms other than text, for examples as drawings or schemes (Sofean & Aras, 2018).

Turner et al. (2005) described heterogeneity within 320 analysed grey literature documents. This included the challenges caused by different document types such as HTML or PDF, different content types such as text or figures or tables, and a general broadness and inconsistency of topics and subject matters (Turner et al., 2005). Other features of unstructured data that cause challenges are colloquialisms, abbreviations, spelling errors and other variations that appear in natural language (Chee et al., 2011; Lee & Uzuner, 2020; Magge, O’ Connor, et al., 2021; Woo et al., 2019).

Chee et al. (2011) describe heterogeneity not within a data source, but between them. Due to different text lengths and/or languages it becomes hard to achieve knowledge– or domain-transfer between sources such as Twitter and online forum entries. This creates a need for developing separate datasets and classifiers for extracting the same type of information, e.g. drugs, from these heterogeneous sources (Chee et al., 2011; Jimeno-Yepes et al., 2015; Rastegar-Mojarad et al., 2016).

###### Complexity, noise, and ambiguity within the data

Complexity is a challenge mostly described within papers that attempted the normalisation of extracted data, due to the large space of possible terms to map to (Batbaatar & Ryu, 2019). Another factor adding on to complexity of tasks is unstructured or irrelevant background information within datasets, or a lack of context within short texts such as tweets (Batbaatar & Ryu, 2019; Gao et al., 2017). Ambiguity also increases complexity, for example when drug name can have multiple synonyms, trade names, or multiple correct labels (Magge, O’ Connor, et al., 2021; Natsiavas et al., 2019). Noise is a concept that generally refers to data being unreliable due to their unstructured and naturally-expressed form, thus causing errors both while labelling gold-standard data and when processing and predicting on new data (Ellendorff et al., 2018; Pasche et al., 2012).

###### Sparsity or imbalance within the data

ML or neural networks architectures require annotated data during the learning process. When training classifiers for forum posts to identify drugs, Chee et al. (2011) noted that some drugs did not have enough mentions in posts in order to train and evaluate robust classifiers, while Arnold et al. (2020) also describe rare entities and unseen data as a problem. Imbalance in the data, when there are more positive training examples for certain types of information, can be another issue that leads to performance drops in under-represented classes (Dai & Wang, 2019; Magge, Tutubalina, et al., 2021).

###### Scalability

Data extraction methods are trained and evaluated on benchmark datasets. However, when deploying them for practical use in real-world scenarios, the amount of data that needs to be processed increases, causing processing times of multiple hours or days and necessitating the use of high-quality hardware and analytics platforms (Keeling et al., 2011; Kravets et al., 2016; Tafti et al., 2017). Ul Haq et al. (2022) discuss that this is a complex task because classification accuracy, time, and versatility need to scale in parallel with the real-world tasks that a system is applied to solve.

###### Corpus availability and cost to generate annotated data

Multiple publications described a lack of publicly corpora and benchmark datasets. This is a commonly described issue for different types of data, including patents (Krishnan et al., 2010) or social media (Yang et al., 2013). It is also mentioned specifically in relation to text extraction and normalization to standardized vocabularies such as the UMLS (Batbaatar & Ryu, 2019)

In the absence of publicly available corpora, researchers are forced to spend money and resources to create their own customized datasets (Chee et al., 2011), which can lead to small datasets with limited usefulness, thus creating the need to adapt models to maximise gain in situations where better performances could be achieved (Shen et al., 2020).

When using labelled gold-standard social-media corpora from sources such as Twitter, copyright and data-availability were described as a challenge (Zhang et al., 2021). Twitter datasets can shrink as tweets become unavailable over time, thus reducing reproducibility and comparability of results obtained at different points in time when using the same corpus (Karisani et al., 2020; Magge, O’ Connor, et al., 2021; Zhang et al., 2021).

However, there exists a vast amount of secondary research that has utilised automated data extraction in practice. These tend to be AI project-specific tools focussing and extracting very targeted and topic-specific information using bespoke methods that are generally not re-usable beyond the original research project. Due to the vast amount of topic-specific automation we did not include these papers in the full-text analysis, but rather tagged them by topic (eg. mental health, COVID-19) and data source (Twitter, news). This supplementary evidence map includes 318 papers. A description of our findings, along with figures and a heat-map is provided in Appendix C, and an interactive version giving access to the papers and their data tags is provided within the SWIFT-Review project in the digital appendix^6^.

## Discussion

The surge in published literature and the dearth of intelligence available is driving the need for more innovative methods to deliver timely secondary research. Fully or semi-automated data extraction may offer a means to make both unstructured and structured data more accessible to those undertaking this type of research. However, conducting any secondary research projects is time-intensive, and often projects themselves are time-sensitive. Therefore, it might not always be feasible to include evidence from lower-quality, grey literature or soft intelligence sources. Fully or semi-automated data extraction can be a way forward, to make unstructured data accessible and facilitate integration into review workflows. However, this works only if data can be automatically identified and extracted using a targeted, well evaluated and evidence-based approach. Further, it remains important that the data being used, whether it is from RCTs or Twitter, are pertinent to the question being asked and the decision being made.

We included 7 end-user tools and 76 published methods papers for data extraction of grey literature and non-peer-reviewed data in this mapping review. There is a broad range of secondary health-related research that could benefit from using grey literature and softer data. Horizon scanning is one use-case, because it utilizes timely, soft sources of information to detect signals of future trends in research and technology (Hines et al., 2019). Similar use-cases for softer and automatically extracted data include the identification of future research topics and protocol formulation. Another potential use-case is the inclusion of rapid analysis of these non-peer-reviewed data sources in the discussion section of systematic reviews, where impact on patients and practitioners, impact on healthcare systems, research gaps in clinical trials or recommendations for the future are discussed.

### Discussion in the context of related literature reviews

Correia et al. (2020) published a narrative review of recent work on data mining in social media content analysis. They discuss papers on automation in the domains of pharmacovigilance and sentiment analysis, most commonly targeting specific drugs and their adverse events, or mental health research questions. These findings correspond to our mapping of the topic-specific literature (see Appendix C), we mapped mental health and sentiment analysis within the top-3 applications. We picked up COVID-19 within the top-3; this is not represented within Correia et al. (2020) due to their publication date in May 2020. They discuss limitations specific to social-media data, such as limitations of reliability of the data when users build online-personas, limitations related to bot-content, limitations when sampling data for analysis, and caveats that people posting online are a small selection from a wider publication and thus samples may not be representative (Correia et al., 2020). A living systematic review of automated data extraction from the highly related field of peer-reviewed health literature currently includes 76 papers, indicating fast-paced advancements in the areas of automatic extraction, normalisation, relation extraction and text summarisation (Schmidt et al., 2021). Their main conclusions are similar to the findings of this review, citing low availability of end-user tools among many published methods of data extraction leading to slow uptake of automation methods in practice, low comparability between evaluation results, and high duplication of research efforts (Schmidt et al., 2021).

### Discussion of important features of extraction tools

Most included papers described methods for extraction of data, with potential features that might be beneficial for future tools. The identified tools discussed accessibility (i.e. as web-application), bulk processing of text, automatically updating data from the web, automatic query expansion, and visualisations as main features. Barriers to integration with other downstream tools was identified as a research gap.

### Discussion of the level of support given by tools and methods

To encourage practical use of the included tools and methods, their underlying NLP methods need to be accessible in the form of usable tools, connected to online data-sources for automatic information retrieval, and well validated. In summary, three different types of evaluation were described in the included publications to validate models, each applied as required by the task and research context:

1. A direct model performance validation, where the proposed model is compared with other published models or algorithms that were trained using the same dataset, ideally with the same train/validation set splits.
2. An adaptability validation, where the proposed model’s evaluation scores are compared with the same model’s scores across different independent datasets that often, but not necessarily, fit the same domain but have different characteristics such as data source, annotation guidelines, or topic-distributions.
3. A practical validation, where the model is used to make predictions on real-life, unlabelled data. This evaluation can be qualitative, in terms of perceived usefulness or trustworthiness of the system as part of a case-study, or comparative in terms of time-saved during screening and data extraction.

### Discussion of practical challenges, research gaps, and caveats as described within the included papers

In the following section, we discuss the broader implications of our analysis, focussing on the limitations associated with using automated data extraction tools and methods in real-world scenarios. These limitations include both the point-of-view of the tool providers, in terms of challenges related to the deployment of usable tools, as well as general challenges caveats relating to user’s lack of trust and further research that is needed when integrating reliable and usable automation in data extraction into real-world research projects.

Natsiavas et al. (2019) described a method and tool design to be used by clinicians at the point of care; noting that integration into already established workflows can be challenging, due to already established routines and information overflow for the clinicians. They noted that having normalised data, facilitating data-sharing, and implementing continuous updates would be helpful, but acknowledge that those features are hard to implement (Natsiavas et al., 2019).

In the real world, tools need to be accessible to users. Costs need to be calculated for hardware and providing computational resources and servers for deployment. This is challenging because it can make it expensive for tools to be live and accessible (Jimeno-Yepes et al., 2015).

Another important issue that prevents the usage of automation methods is a lack of trust in the reliability of tools and methods, concerning for example trust into the sources of the information or a lack of high-performing classifiers that can provide adequate performance (Keeling et al., 2011; Magge, Tutubalina, et al., 2021; Valentin et al., 2021). Many of the included papers used their own datasets for training and evaluation, or completed their evaluation using different evaluation metrics. This severely limits the comparability of approaches and cannot provide us with definite answers on how trustworthy or reliable tools are. Whenever methods were tested in real-world scenarios, these tests were usually small and unmeaningful as the process of using the automation approach was not directly compared with a fully manual analyses on the same dataset. Outcomes such as time-saved, or number of relevant records discovered by each method, were rarely assessed.

Non-scientific or grey literature data from social media, patents, or similar sources are often expressed in languages other than English. This means that problems around sparsity, data imbalance, and lack of availability are exacerbated, as they make it harder to obtain good representations of the language and tasks that need to be achieved. A potential solution to this problem is the usage of automatic translation software. With the advent of neural networks in NLP the performance of machine-translation algorithms has steadily improved over the past years (Wang et al., 2022), and a selection of free or paid-for APIs such as DeepL^7^ and Google Translate^8,9^ are available to process information in various formats.

A potential practical challenge we noted is the evaluation of a tool or model on data that has previously been seen in training. This issue should not arise when training and evaluating on one dataset that has been correctly split into train and test data, but it may arise when multiple datasets are created from the same source and then subsequently used as additional evaluation sets. For example, ClinicalTrials.gov is a frequent source of data described in multiple papers and datasets (Goodwin et al., 2018; Patel & Cimino, 2007; Pradhan et al., 2019; Smalheiser & Holt, 2022; Tian et al., 2021) and thus caution needs to be exercised when using or evaluating across datasets that are available from related research projects.

### Recommendations for tool development

In summary, researchers or companies looking to develop automated data extraction tools should consider the financial implications for tool development and deployment, and the scalability of their automation methods to estimate hardware needs and running costs in the long-term. They should also carefully assess user needs and interoperability of the proposed tool with other down– or upstream tools used in literature analysis or in clinical practice. These considerations determine the complexity of their proposed tool and should be key to the planning, execution, and evaluation stages of the final tool.

Unfortunately, this does not guarantee user-acceptance, and significant risks remain when investing research time and money into tool development. A lack of trust in the tool and/or its underlying automation methods may still lead to a lower-than-expected uptake, at which point the effort of maintaining tools is too high and may not be worth it.

To increase acceptance, transparent large-scale testing and comparisons on different datasets may be needed, in conjunction with early engagement with the research community during the design phase and later via publications in peer-reviewed journals. As discussed in the previous sections, a comprehensive evaluation includes comparing and contrasting one automation model with other models using the same datasets, applying the model to multiple datasets with different characteristics to simulate different real-world projects, and running large-scale practical evaluations on real-world projects without any pre-defined gold-standard data, to measure outcomes important to researchers who conduct literature reviews. These outcomes could be time-savings between automation-supported and manual processes, number of records missed or gained through automation, and usability and integrability of the tool into established workflows and methodologies. None of the included papers mentioned marketing, to increase awareness and public knowledge about tools.

### Limitations

The scope of this mapping review is intentionally broad. A vast amount of literature exists in the intersection between automatic data extraction and health-related evidence/data in the public domain. We aimed to be systematic in capturing the relevant literature during the search but limited the inclusion criteria to papers that extract general-purpose text (as opposed to including topic-specific analyses). To mitigate this limitation, we have separated and tagged all topic-specific papers as part of a separate evidence map based on title/abstract information of 318 papers. We presented an abbreviated version of these results within Appendix C.

We extracted evaluation scores for included papers and discussed model performances for tasks of differing complexity but did not directly compare the performance of any methods discussed in this review. We avoided drawing conclusions on the ‘best’ tools or methods available; this was not our aim. However, had we sought to do this feasibility of such an exercise would have been hindered by the usage of different datasets and different methods of evaluations across the papers. Further in-depth case-studies that include implementation and direct comparisons of some of these methods are reserved for future work.

## Conclusion

This review summarises current knowledge about functionalities, data sources and performance of published methods and tools to automate data extraction of grey literature and softer intelligence related to healthcare. We performed a detailed analysis of key strengths and weaknesses of 7 end-user tools and 76 methods papers, and the level of support they provide. We collected information about barriers in implementing automation in practice, and a summary of caveats and experiences from using automatically mined and extracted data in real world projects. Overall availability of code, data, and implementation of methods into accessible end-user tools was poor, suggesting that the field of automating grey-literature mining suffers from high duplication of research efforts, and at the same time low uptake of the few tools and methods that are available.

## Highlights

This is the first review of automated data extraction from health-related grey literature and soft intelligence; to automate horizon scans, HTAs, evidence maps or other secondary literature reviews. It includes 84 tools and methods papers mining information from health-related news, patents, websites, trial registers, fora, or social media.

We discuss relevant end-user features of tools, types of extracted data and text such as ‘disease’ or ‘outcome’, evaluation metrics and results, practical implications of usage, research gaps and barriers to development and deployment of automation methods in this field in practice.

This review provides a detailed insight into automated classification, mining, and normalisation of data from grey literature and softer intelligence. We tagged, mapped, and shared all results to enable both data scientists and researcher with health-related research background to easily filter and access all included papers.

## Supporting information

Exclusion Reasons

## Data Availability

All data produced are available online at:
Schmidt, Lena, 2023, “Automated data extraction of unstructured and grey literature data in health research: a mapping review of the current research literature”, https://doi.org/10.7910/DVN/7N2YWZ, Harvard Dataverse

## Acknowledgements

We would like to thank Chris Marshall for giving very valuable feedback on the methodology of this paper during protocol development and for providing edits and feedback to an earlier draft.

We would also like to thank Fiona Beyer for her feedback on the search strategies and her recommendations regarding the databases to search for this review.

## Data availability statement

All references included on full text and for the separate map of titles and abstracts, together with their tags and extracted data, are available in the form of SWIFT-Review project files in the digital appendix of this paper. We also created a repository on Harvard Dataverse to host the project files, it is available here:

Schmidt, Lena, 2023, “Automated data extraction of unstructured and grey literature data in health research: a mapping review of the current research literature”, https://doi.org/10.7910/DVN/7N2YWZ, Harvard Dataverse

## Contributions

LS: Conceptualization, Methodology, development of search, investigation, formal analysis, visualization, writing (original draft)

SM: Methodology, investigation, writing (review & editing)

NM: Methodology, investigation, writing (review & editing), supervision

JB: Methodology, writing (review & editing), supervision

DC: Conceptualization, Methodology, writing (review & editing), supervision

## Other statements

- ethics approval statement: Not applicable
- patient consent statement: Not applicable
- permission to reproduce material from other sources: Not applicable
- clinical trial registration: Not applicable

## Footnotes

1 https://www.merriam-webster.com/words-at-play/words-were-watching-infodemic-meaning

2 https://www.python.org/doc/versions/

3 https://www.java.com/releases/

4 https://cran.r-project.org/web/packages/medrxivr/index.html

5 https://live.european-language-grid.eu/catalogue/corpus/5090

6 https://doi.org/10.7910/DVN/7N2YWZ

7 https://www.deepl.com/pro-api?cta=header-pro-api

8 https://cloud.google.com/translate

9 https://py-googletrans.readthedocs.io/en/latest/

## References

1. Acosta-Urigüen, M.-I., Arias, B., & Orellana, M. (2020). Text Mining Techniques Implemented to Extract Data from Transit Events in Twitter: A Systematic Literature Review. In G. Rodriguez Morales, E. R. Fonseca, C. J. P. Salgado, P. Pérez-Gosende, M. Orellana Cordero, & S. Berrezueta, Information and Communication Technologies Cham.

2. Alvaro, N., Miyao, Y., & Collier, N. (2017). TwiMed: Twitter and PubMed Comparable Corpus of Drugs, Diseases, Symptoms, and Their Relations. JMIR Public Health Surveill, 3(2), e24. https://doi.org/10.2196/publichealth.6396

3. Arnold, S., Van Aken, B., Grundmann, P., Gers, F. A., & Löser, A. (2020). Learning Contextualized Document Representations for Healthcare Answer Retrieval.

4. Atal, I., Zeitoun, J. D., Névéol, A., Ravaud, P., Porcher, R., & Trinquart, L. (2016). Automatic classification of registered clinical trials towards the Global Burden of Diseases taxonomy of diseases and injuries. BMC Bioinformatics, 17(1), 392. https://doi.org/10.1186/s12859-016-1247-7

5. Avasarala, V., & Bonissone, P. (2012). iPresage: An innovative patent landscaping tool.

6. Batbaatar, E., & Ryu, K. H. (2019). Ontology-Based Healthcare Named Entity Recognition from Twitter Messages Using a Recurrent Neural Network Approach. Int J Environ Res Public Health, 16(19). https://doi.org/10.3390/ijerph16193628

7. Blei, D. M., Ng, A. Y., & Jordan, M. I. (2003). Latent dirichlet allocation. J. Mach. Learn. Res., 3(null), 993– 1022.

8. Blumenfeld, P., Pfeffer, R. M., Symon, Z., Den, R. B., Dicker, A. P., Raben, D., & Lawrence, Y. R. (2014). The lag time in initiating clinical testing of new drugs in combination with radiation therapy, a significant barrier to progress? British Journal of Cancer, 111(7), 1305–1309. https://doi.org/10.1038/bjc.2014.448

9. Cabanac, G., Oikonomidi, T., & Boutron, I. (2021). Day-to-day discovery of preprint–publication links. Scientometrics, 126(6), 5285–5304. https://doi.org/10.1007/s11192-021-03900-7

10. Chee, B. W., Berlin, R., & Schatz, B. (2011). Predicting adverse drug events from personal health messages [Article]. AMIA Annual Symposium proceedings / AMIA Symposium. AMIA Symposium, 2011, 217–226. https://www.scopus.com/inward/record.uri?eid=2-s2.0-84863556111&partnerID=40&md5=72d17f7324c0865344858656d85b7506

11. Chen, Y., Zhou, C., Li, T., Wu, H., Zhao, X., Ye, K., & Liao, J. (2019). Named entity recognition from Chinese adverse drug event reports with lexical feature based BiLSTM-CRF and tri-training [Article]. Journal of Biomedical Informatics, 96, Article 103252. https://doi.org/10.1016/j.jbi.2019.103252

12. Correia, R. B., Wood, I. B., Bollen, J., & Rocha, L. M. (2020). Mining Social Media Data for Biomedical Signals and Health-Related Behavior. Annu Rev Biomed Data Sci, 3, 433–458. https://doi.org/10.1146/annurev-biodatasci-030320-040844

13. Dai, H. J., & Wang, C. K. (2019). Classifying adverse drug reactions from imbalanced twitter data. Int J Med Inform, 129, 122–132. https://doi.org/10.1016/j.ijmedinf.2019.05.017

14. Daniel, C., & Dutta, K. (2018). Automated generation of latent topics on emerging technologies from YouTube video content.

15. DeYoung, J., Beltagy, I., van Zuylen, M., Kuehl, B., & Wang, L. L. (2021). MŜ2: A Dataset for Multi-Document Summarization of Medical Studies. ArXiv. https://doi.org/https://doi.org/10.48550/arXiv.2104.06486

16. Ellendorff, T., Cornelius, J., Gordon, H., Colic, N., & Rinaldi, F. (2018). UZH@SMM4H: System Descriptions. https://doi.org/10.18653/v1/W18-5916

17. Fan, B., Fan, W., Smith, C., & Garner, H. (2020). Adverse drug event detection and extraction from open data: A deep learning approach [Article]. Information Processing and Management, 57(1), Article 102131. https://doi.org/10.1016/j.ipm.2019.102131

18. Gao, J., Liu, N., Lawley, M., & Hu, X. (2017). An Interpretable Classification Framework for Information Extraction from Online Healthcare Forums [Article]. Journal of Healthcare Engineering, 2017, Article 2460174. https://doi.org/10.1155/2017/2460174

19. Goodman, C. S., & Church, F. (2004). HTA 101 INTRODUCTION TO HEALTH TECHNOLOGY ASSESSMENT.

20. Goodwin, T. R., Skinner, M. A., & Harabagiu, S. M. (2018). Automatically Linking Registered Clinical Trials to their Published Results with Deep Highway Networks. AMIA Jt Summits Transl Sci Proc, 2017, 54–63.

21. Grabar, N., & Hamon, T. (2014). Automatic extraction of layman names for technical medical terms.

81. Guo, H., Na, X., & Li, J. (2017). Automatically Identifying Topics of Consumer Health Questions in Chinese. Stud Health Technol Inform, 245, 388–392.

22. Guo, Y., Ge, Y., Yang, Y. C., Al-Garadi, M. A., & Sarker, A. (2022). Comparison of pretraining models and strategies for health-related social media text classification. https://doi.org/10.1101/2021.09.28.21264253

23. Haddaway, N. R., Page, M. J., Pritchard, C. C., & McGuinness, L. A. (2022). PRISMA2020: An R package and Shiny app for producing PRISMA 2020-compliant flow diagrams, with interactivity for optimised digital transparency and Open Synthesis [https://doi.org/10.1002/cl2.1230]. Campbell Systematic Reviews, 18(2), e1230. https://doi.org/https://doi.org/10.1002/cl2.1230

24. Halim, C., Wicaksono, A. F., & Adriani, M. (2018). Extracting disease-symptom relationships from health question and answer forum.

25. Hariprasad, S., Xue-wen, C., & Bo, L. (2015). Ontology-Based Visualization of Healthcare Data Mined from Online Healthcare Forums. https://doi.org/10.1109/ICHI.2015.46

26. Hines, P., Hiu Yu, L., Guy, R. H., Brand, A., & Papaluca-Amati, M. (2019). Scanning the horizon: a systematic literature review of methodologies. BMJ Open, 9(5), e026764. https://doi.org/10.1136/bmjopen-2018-026764

27. Hopewell, S., McDonald, S., Clarke, M., & Egger, M. (2007). Grey literature in meta-analyses of randomized trials of health care interventions. Cochrane Database Syst Rev, 2007(2), Mr000010. https://doi.org/10.1002/14651858.MR000010.pub3

28. Howard, B., Phillips, J., Tandon, A., Maharana, A., Elmore, R., Mav, D., Sedykh, A., Thayer, K., Merrick, A., Walker, V., Rooney, A., & Shah, R. (2020). SWIFT-Active Screener: Accelerated document screening through active learning and integrated recall estimation. Environment International, 138, 105623. https://doi.org/https://doi.org/10.1016/j.envint.2020.105623 10.3389/fdgth.2020.592237

29. Howard, B. E., Phillips, J., Miller, K., Tandon, A., Mav, D., Shah, M. R., Holmgren, S., Pelch, K. E., Walker, V., Rooney, A. A., Macleod, M., Shah, R. R., & Thayer, K. (2016). SWIFT-Review: a text-mining workbench for systematic review. Syst Rev, 5(1), 87. https://doi.org/10.1186/s13643-016-0263-z

30. Jimeno-Yepes, A., MacKinlay, A., Han, B., & Chen, Q. (2015). Identifying Diseases, Drugs, and Symptoms in Twitter. Stud Health Technol Inform, 216, 643–647.

31. Jonnalagadda, S. R., Goyal, P., & Huffman, M. D. (2015). Automating data extraction in systematic reviews: a systematic review. Systematic Reviews, 4(1). https://doi.org/10.1186/s13643-015-0066-7

32. Karimi, S., Metke-Jimenez, A., Kemp, M., & Wang, C. (2015). Cadec: A corpus of adverse drug event annotations. J Biomed Inform, 55, 73–81. https://doi.org/10.1016/j.jbi.2015.03.010

33. Karisani, P., Ho, J., & Agichtein, E. (2020). Domain-Guided Task Decomposition with Self-Training for Detecting Personal Events in Social Media. https://export.arxiv.org/abs/2004.10201

34. Keeling, J. W., Turner, A. M., Allen, E. E., Rowe, S. A., Merrill, J. A., Liddy, E. D., & Turtle, H. R. (2011). Development and evaluation of a prototype search engine to meet public health information needs. AMIA Annu Symp Proc, 2011, 693–700.

35. Kocijan, K., Kurolt, S., & Mijić, L. (2020). Building croatian medical dictionary from medical corpus [Article]. Rasprave Instituta za Hrvatski Jezik i Jezikoslovlje, 46(2), 765–782. https://doi.org/10.31724/RIHJJ.46.2.17

36. Korkontzelos, I., Nikfarjam, A., Shardlow, M., Sarker, A., Ananiadou, S., & Gonzalez, G. H. (2016). Analysis of the effect of sentiment analysis on extracting adverse drug reactions from tweets and forum posts [Article]. Journal of Biomedical Informatics, 62, 148–158. https://doi.org/10.1016/j.jbi.2016.06.007

37. Kravets, A. G., Korobkin, D. M., & Dykov, M. A. (2016). E-patent examiner: Two-steps approach for patents prior-art retrieval.

38. Krishnan, A., Cardenas, A. F., & Springer, D. (2010). Search for patents using treatment and causal relationships.

39. Lauvrak, V., Arentz-Hansen, H., & Di Bidino, R. (2020). Recommendations for Horizon Scanning, Topic Identification, Selection and Prioritisation for European Cooperation on Health Technology Assessment. EUnetHTA WP4 Deliverable 4.10. https://www.eunethta.eu/wp-content/uploads/2020/04/200305-EUnetHTA-WP4-Deliverable-4.10-TISP-recommendations-final-version-1.pdf

40. Lee, K., & Uzuner, Ö. (2020). Normalizing Adverse Events using Recurrent Neural Networks with Attention. AMIA Jt Summits Transl Sci Proc, 2020, 345–354.

41. Lefebvre, C., Glanville, J., Briscoe, S., Littlewood, A., Marshall, C., Metzendorf, M.-I., Noel-Storr, A., Rader, T., Shokraneh, F., Thomas, J., Wieland, L. S., & on behalf of the Cochrane Information Retrieval Methods, G. (2019). Searching for and selecting studies. In Cochrane Handbook for Systematic Reviews of Interventions (pp. 67–107). https://doi.org/https://doi.org/10.1002/9781119536604.ch4

42. Liu, S., Bourgeois, F. T., & Dunn, A. G. (2022). Identifying unreported links between ClinicalTrials.gov trial registrations and their published results [https://doi.org/10.1002/jrsm.1545]. Research Synthesis Methods, 13(3), 342–352. https://doi.org/https://doi.org/10.1002/jrsm.1545

43. Magge, A., O’ Connor, K., Scotch, M., & Gonzalez-Hernandez, G. (2021). SEED: Symptom Extraction from English Social Media Posts using Deep Learning and Transfer Learning. medRxiv. https://doi.org/10.1101/2021.02.09.21251454

44. Magge, A., Tutubalina, E., Miftahutdinov, Z., Alimova, I., Dirkson, A., Verberne, S., Weissenbacher, D., & Gonzalez-Hernandez, G. (2021). DeepADEMiner: a deep learning pharmacovigilance pipeline for extraction and normalization of adverse drug event mentions on Twitter. J Am Med Inform Assoc, 28(10), 2184–2192. https://doi.org/10.1093/jamia/ocab114

45. McGuinness, L. A., & Schmidt, L. (2020). medrxivr: Accessing and searching medRxiv and bioRxiv preprint data in R. The Journal of Open Source Software, 5(54). https://doi.org/https://doi.org/10.21105/joss.02651

46. Miller, D. M., & Shalhout, S. Z. (2021). GENETEX—a GENomics Report TEXt mining R package and Shiny application designed to capture real-world clinico-genomic data. JAMIA Open, 4(3), ooab082. https://doi.org/10.1093/jamiaopen/ooab082

47. Morris, Z. S., Wooding, S., & Grant, J. (2011). The answer is 17 years, what is the question: understanding time lags in translational research. J R Soc Med, 104(12), 510–520. https://doi.org/10.1258/jrsm.2011.110180

48. Natsiavas, P., Jaulent, M. C., & Koutkias, V. (2019). A Knowledge-Based Platform for Assessing Potential Adverse Drug Reactions at the Point of Care: User Requirements and Design. Stud Health Technol Inform, 264, 1007–1011. https://doi.org/10.3233/shti190376

49. O’Mara-Eves, A., Thomas, J., McNaught, J., Miwa, M., & Ananiadou, S. (2015). Using text mining for study identification in systematic reviews: a systematic review of current approaches. Systematic Reviews, 4(1). https://doi.org/10.1186/2046-4053-4-5

50. Paez, A. (2017). Grey literature: An important resource in systematic reviews. J Evid Based Med. https://doi.org/10.1111/jebm.12265

51. Page, M. J., McKenzie, J. E., Bossuyt, P. M., Boutron, I., Hoffmann, T. C., Mulrow, C. D., Shamseer, L., Tetzlaff, J. M., Akl, E. A., Brennan, S. E., Chou, R., Glanville, J., Grimshaw, J. M., Hróbjartsson, A., Lalu, M. M., Li, T., Loder, E. W., Mayo-Wilson, E., McDonald, S., Moher, D. (2021). The PRISMA 2020 statement: an updated guideline for reporting systematic reviews. BMJ, 372, n71. https://doi.org/10.1136/bmj.n71

52. Pasche, E., Gobeill, J., Teodoro, D., Gaudinat, A., Vishnyakova, D., Lovis, C., & Ruch, P. (2012). A user-friendly tool for medical-related patent retrieval. Stud Health Technol Inform, 174, 121–125.

53. Patel, C. O., & Cimino, J. J. (2007). Semantic query generation from eligibility criteria in clinical trials. AMIA Annu Symp Proc, 1070.

54. Pradhan, R., Hoaglin, D. C., Cornell, M., Liu, W., Wang, V., & Yu, H. (2019). Automatic extraction of quantitative data from ClinicalTrials.gov to conduct meta-analyses. J Clin Epidemiol, 105, 92–100. https://doi.org/10.1016/j.jclinepi.2018.08.023

55. Rabatel, J., Arsevska, E., & Roche, M. (2019). PADI-web corpus: Labeled textual data in animal health domain. Data Brief, 22, 643–646. https://doi.org/10.1016/j.dib.2018.12.063

56. Rastegar-Mojarad, M., Liu, H., & Nambisan, P. (2016). Using Social Media Data to Identify Potential Candidates for Drug Repurposing: A Feasibility Study. JMIR Res Protoc, 5(2). https://doi.org/10.2196/resprot.5621

57. Rezaei, Z., Ebrahimpour-Komleh, H., Eslami, B., Chavoshinejad, R., & Totonchi, M. (2020). Adverse Drug Reaction Detection in Social Media by Deepm Learning Methods. Cell J, 22(3), 319–324. https://doi.org/10.22074/cellj.2020.6615

58. Saha, S., Das, S., Khurana, P., & Srihari, R. (2020). Autobots Ensemble: Identifying and Extracting Adverse Drug Reaction from Tweets Using Transformer Based Pipelines. https://aclanthology.org/2020.smm4h-1.16

59. Schmidt, L., Olorisade, B. K., McGuinness, L. A., Thomas, J., & Higgins, J. P. T. (2021). Data extraction methods for systematic review (semi)automation: A living systematic review. F1000Research, 10, 401.

60. Shen, C., Lin, H., Li, Z., Chu, Y., & Yang, Z. (2020). A Graph-boosted Framework for Adverse Drug Event Detection on Twitter.

61. Singh, L., Bode, L., Budak, C., Kawintiranon, K., Padden, C., & Vraga, E. (2020). Understanding high– and low-quality URL Sharing on COVID-19 Twitter streams. Journal of Computational Social Science, 3(2), 343–366. https://doi.org/10.1007/s42001-020-00093-6

62. Smalheiser, N. R., & Holt, A. W. (2022). A web-based tool for automatically linking clinical trials to their publications. J Am Med Inform Assoc. https://doi.org/10.1093/jamia/ocab290

63. Sofean, M., & Aras, H. (2018). Technological areas detection and clustering for large-scale of patent texts.

64. Stenetorp, P., Pyysalo, S., Topić, G., Ohta, T., Ananiadou, S., & Tsujii, J. i. (2012, April). brat: a Web-based Tool for NLP-Assisted Text Annotation.Proceedings of the Demonstrations at the 13th Conference of the European Chapter of the Association for Computational Linguistics Avignon, France.

65. Tafti, A. P., Badger, J., LaRose, E., Shirzadi, E., Mahnke, A., Mayer, J., Ye, Z., Page, D., & Peissig, P. (2017). Adverse Drug Event Discovery Using Biomedical Literature: A Big Data Neural Network Adventure. JMIR Med Inform, 5(4). https://doi.org/10.2196/medinform.9170

66. Tian, S., Erdengasileng, A., Yang, X., Guo, Y., Wu, Y., Zhang, J., Bian, J., & He, Z. (2021). Transformer-based named entity recognition for parsing clinical trial eligibility criteria.

67. Turner, A. M., Liddy, E. D., Bradley, J., & Wheatley, J. A. (2005). Modeling public health interventions for improved access to the gray literature. J Med Libr Assoc, 93(4), 487–494.

68. Ul Haq, H., Kocaman, V., & Talby, D. (2022). Mining Adverse Drug Reactions from Unstructured Mediums at Scale. ArXiv. https://doi.org/https://doi.org/10.48550/arXiv.2201.01405

69. Valentin, S., Arsevska, E., Rabatel, J., Falala, S., Mercier, A., Lancelot, R., & Roche, M. (2021). PADI-web 3.0: A new framework for extracting and disseminating fine-grained information from the news for animal disease surveillance. One Health, 13, 100357. https://doi.org/10.1016/j.onehlt.2021.100357

70. Van Norman, G. A. (2016). Drugs, Devices, and the FDA: Part 2: An Overview of Approval Processes: FDA Approval of Medical Devices. JACC: Basic to Translational Science, 1(4), 277–287. https://doi.org/https://doi.org/10.1016/j.jacbts.2016.03.009

71. Viviani, M., & Pasi, G. (2017). Credibility in social media: opinions, news, and health information—a survey [https://doi.org/10.1002/widm.1209]. WIREs Data Mining and Knowledge Discovery, 7(5), e1209. https://doi.org/https://doi.org/10.1002/widm.1209

72. Wang, H., Wu, H., He, Z., Huang, L., & Church, K. W. (2022). Progress in Machine Translation. Engineering, 18, 143–153. https://doi.org/https://doi.org/10.1016/j.eng.2021.03.023

73. WHO. (2021). Health Technology Assessment Survey 2020/21 – Main Findings. WHO. https://www.who.int/data/stories/health-technology-assessment-a-visual-summary

74. Woo, H. G., Yeom, J., & Lee, C. (2019). Screening early stage ideas in technology development processes: a text mining and k-nearest neighbours approach using patent information [Article]. Technology Analysis and Strategic Management, 31(5), 532–545. https://doi.org/10.1080/09537325.2018.1523386

75. Xiao, C., Choi, E., & Sun, J. (2018). Opportunities and challenges in developing deep learning models using electronic health records data: a systematic review. Journal of the American Medical Informatics Association, 25(10), 1419–1428. https://doi.org/10.1093/jamia/ocy068

76. Yang, M., Wang, X., & Kiang, M. (2013). Identification of consumer Adverse Drug Reaction messages on social media.

77. Zhang, T., Lin, H., Xu, B., Yang, L., Wang, J., & Duan, X. (2021). Adversarial neural network with sentiment-aware attention for detecting adverse drug reactions. J Biomed Inform, 123, 103896. https://doi.org/10.1016/j.jbi.2021.103896

78. Zhao, B., Xu, S., Lin, S., Luo, X., & Duan, L. (2016). A new visual navigation system for exploring biomedical Open Educational Resource (OER) videos. J Am Med Inform Assoc, 23(e1). https://doi.org/10.1093/jamia/ocv123

79. Zhao, S., Jiang, M., Yuan, Q., Qin, B., Liu, T., & Zhai, C. (2017). ContextCare: Incorporating contextual information networks to representation learning on medical forum data.

80. Zhou, Y., Dong, F., Liu, Y., & Ran, L. (2021). A deep learning framework to early identify emerging technologies in large-scale outlier patents: an empirical study of CNC machine tool [Article]. Scientometrics, 126(2), 969–994. https://doi.org/10.1007/s11192-020-03797-8

