## Appendices for "Automated data extraction of unstructured grey literature in health research: a mapping review of the current research literature"

### Appendix A: Full research questions and items for data extraction

**RQ1: What are the most important features of an extraction tool or method to support health-related research?**

Data extraction items for this research question:

1. List of relevant features and functionalities, as described in each paper.
2. From which source are data retrieved and extracted? Answers could be, for example, ‘Twitter’ or ‘GoogleNews’.
3. Which types of data are extracted?

**RQ2: What level of support can existing tools or methods provide to expedite evidence extraction in health-related research?**

Data extraction items for this research question:

1. Does the reference describe an end-user tool?
2. To which extend is data extraction automated? Exemplary answer can be ‘prioritisation of likely content’, ‘data mining’ or ‘extraction and normalization to standardized vocabulary’.
3. Description of the implementation and evaluation of the tool’s performance. Where possible, we will extract evaluation scores such as recall, precision, or F1.
4. Description of the tool’s integration into current review workflow, as described within each reference.

**RQ3: What are practical challenges and research gaps that constitute barriers to the development and deployment of data extraction tools and methods?**

Data extraction items for this research question:

1. Challenges or barriers that hinder the development or deployment of new tools, as described in each paper.
2. Challenges or caveats for the usage of this tool in real-world research projects, as described in each paper.
3. Research gaps and other practical challenges, as described in each paper.

### Appendix B: Glossary

Algorithm: An algorithm is a clearly defined process with step-wise instructions to solve a particular task. It can be, for example, a natural language processing method to summarise text. In this example, steps might be taken such as identification of important pieces of information, merging them, and creating new and short grammatically correct sentences.

Data mining: Data mining is the process ofidentifying data, often sentences or named entities of a specific type, from multiple documents. The number of documents can be large, and they do not need to be confined to a specific topic. The objective of data mining is to find, and save, all entities within a document that are relevant to a specific class. Approaches commonly summarised under the term data mining are for example named-entity recognition, sentence-classification, or extractive question answering.

Document classification: In the field of Natural Language Processing (NLP), document classification is also referred to as ’sequence classification’. The aim of this task is the categorization of sentences, paragraphs or longer sequences of text. Classification is usually binary or multi-class, assigning documents to a clearly defined decision-space. An example from the field of systematic review automation for binary document classification is classifying abstracts into in- and excluded categories. An example for multi-class classification is sentence
into classification into population, intervention/control, outcomes.

Entity: A short piece of text that can be categorized as belonging to a specific class. For example, the term ’acetylsalicylic acid’ is an entity of the class ’Drug’.

Entity normalization: Entity normalization is the process of grouping entities that refer to the same concept, and matching them to a controlled vocabulary. For example, after named-entity recognition, a document could contain the drug entities ’acetylsalicylic acid’ and ’Acetysal’. Those terms could be normalised to the MeSH term D001241: ’Aspirin’.

### Appendix C: Evidence-mapping on title and abstract level

We applied topic and data tags to 318 titles and abstracts that were not part of the full-text analysis because they described bespoke, topic-specific automation methods. Those methods were applied in practice to answer specific research questions but are of limited value beyond that scope. The full set of interactive results, including tagged papers, is available in the form of a SWIFT-Review project which can be found in the digital appendix of this paper. Figure 7 shows the heatmap of the intersections between topics and data sources. Currently, the most prevalent source of data tagged in health-related research is Twitter (n=122), followed by content from health-related fora, such as Ask a Patient or MedHelp^[1]^ (n=56). Mental health and Covid-19 are the most prevalent health topics in the literature. We tagged n=20 papers on the intersection of Twitter and Covid-19, and a further n=18 papers for the more general topic of disease outbreaks, which includes research on flu, ebola, and epidemics in general.

Interestingly, we did not find much intersecting research for Covid-19 on internet fora, most data about Covid-19 related research are derived from Twitter, followed by news (n=6) and search engines (n=5). In contrast to Covid-19 research, mental health research is commonly conducted on data from fora (n=11). However, even in this comparison, Twitter was the most prevalent source of data (n=19), followed by the generic tag ‘social media’ which was applied to papers that indicated social media as data source without further specifying a more detailed source of data in the abstract (n=13). Overall, mental health had the highest numbers of papers in the topic-space (n=56), but the research topic of Covid-19 (n=54) has been applied to a larger variety of data sources.

In the following, we will take a closer look at the topic and data tags separately.

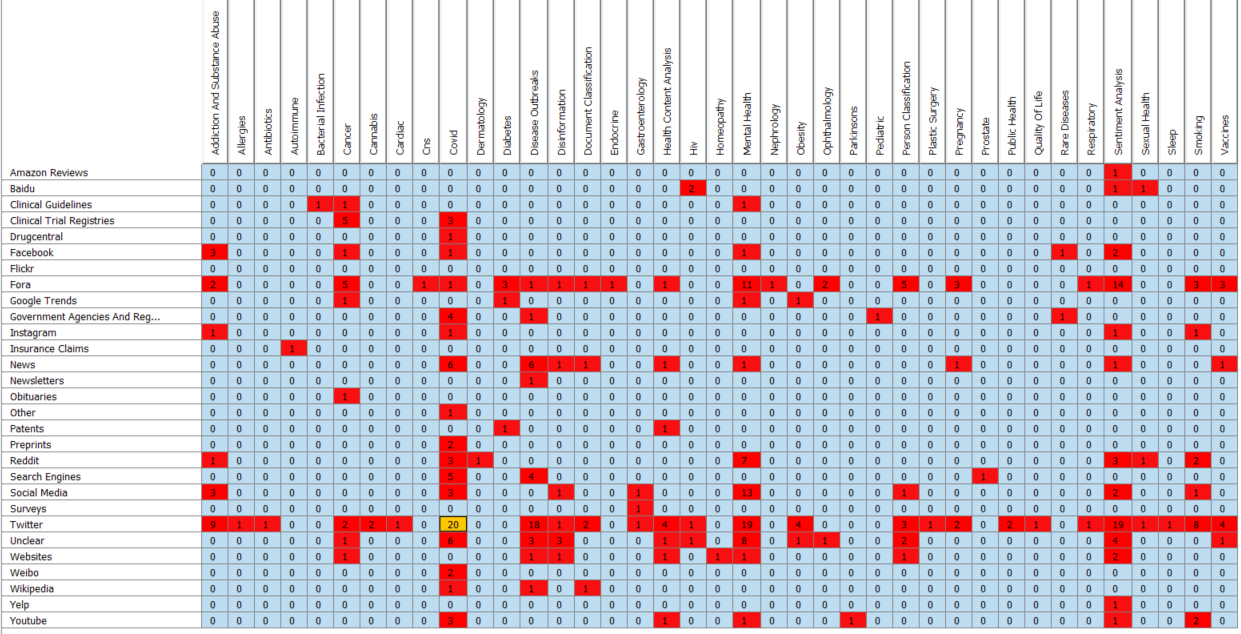

Figure 7: Heatmap showing intersections between the research topic and data sources described in topic-specific titles and abstracts.

Figure 7 visualises the breakdown and proportion of contribution of different data sources to 318 papers tagged in this supplementary evidence map. Twitter and online fora were the two major contributors within our 28 different data sources. Several papers just generally mentioned social media as data source or were unclear about the actual data sources (n=25 and n=31). In general, we observed a large variety of different data media within social network data, including text data (Reddit n=17, Facebook n=8), video data (Youtube n=8) or pictures (Instagram n=3). Besides social-media-related data, news (n=17), search engines (n=10), clinical trial registries (n=8), and government agencies and registers (n=7) were prevalent sources of data.

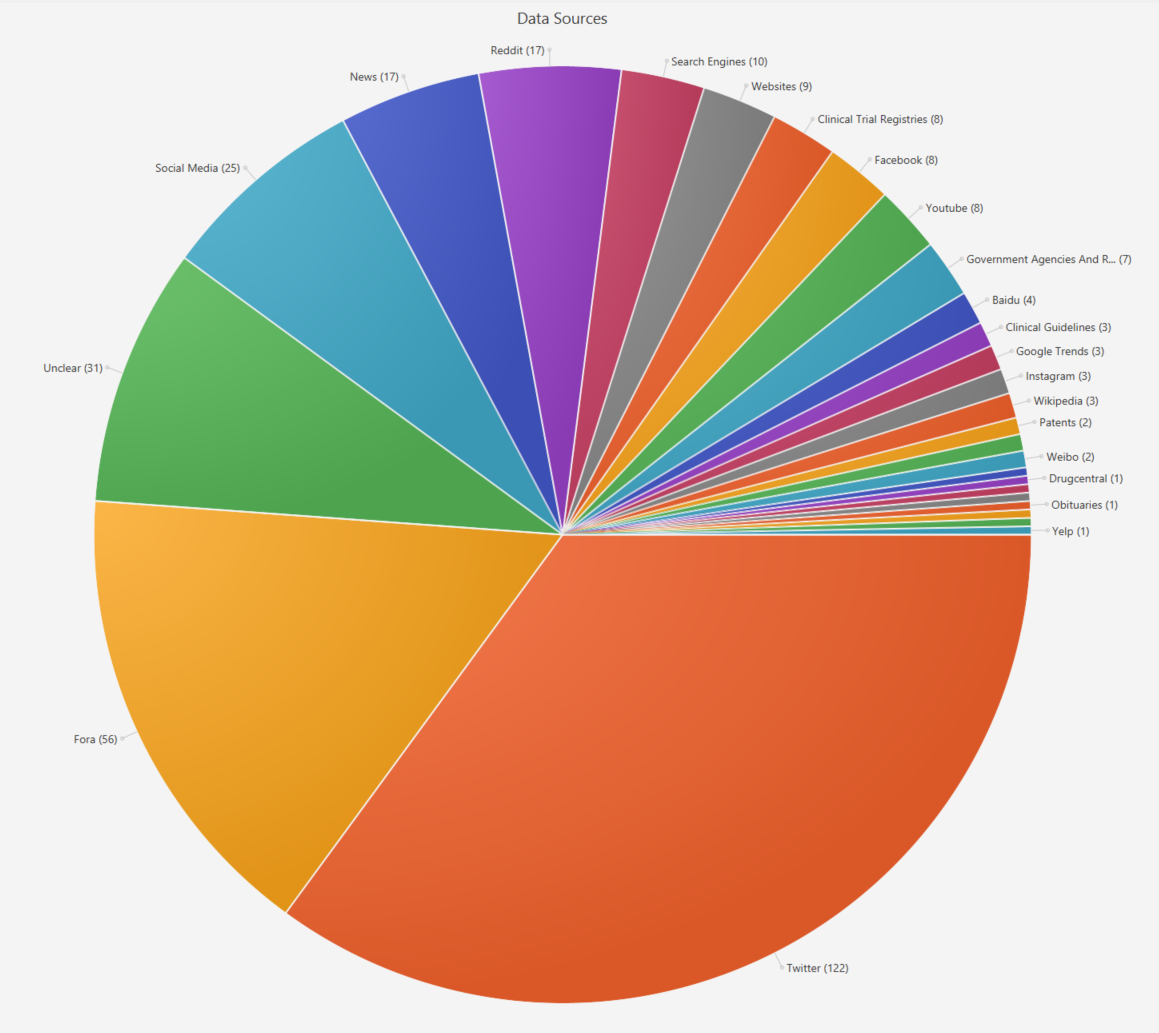

Figure 8: Sources of data in the tagged titles and abstracts. We aimed to make tags as granular as possible, tagging references using different social media data source separately when possible.

Figure 8 shows the most prevalent health-topics studied in the tagged titles and abstracts. Mental health, with 60 references, is the most widely tagged topic. However, we tagged Covid-19 separately to the generic tag for infectious diseases. When combined, those two topics would yield more papers than mental health alone. In the context of this tag category it was possible to apply more than one topic tag to each reference. Especially in the generic category for sentiment analysis, which was tagged n=47 times, we applied co-tags in vaccine research, drug-research, or tobacco and vaping-related research. There was a wide variety of research topic, spanning rare diseases (n=1), multiple medical sub-domains, all the way to very prevalent diseases such as Covid-19 and common issues such as substance abuse (n=19) and obesity (n=6).

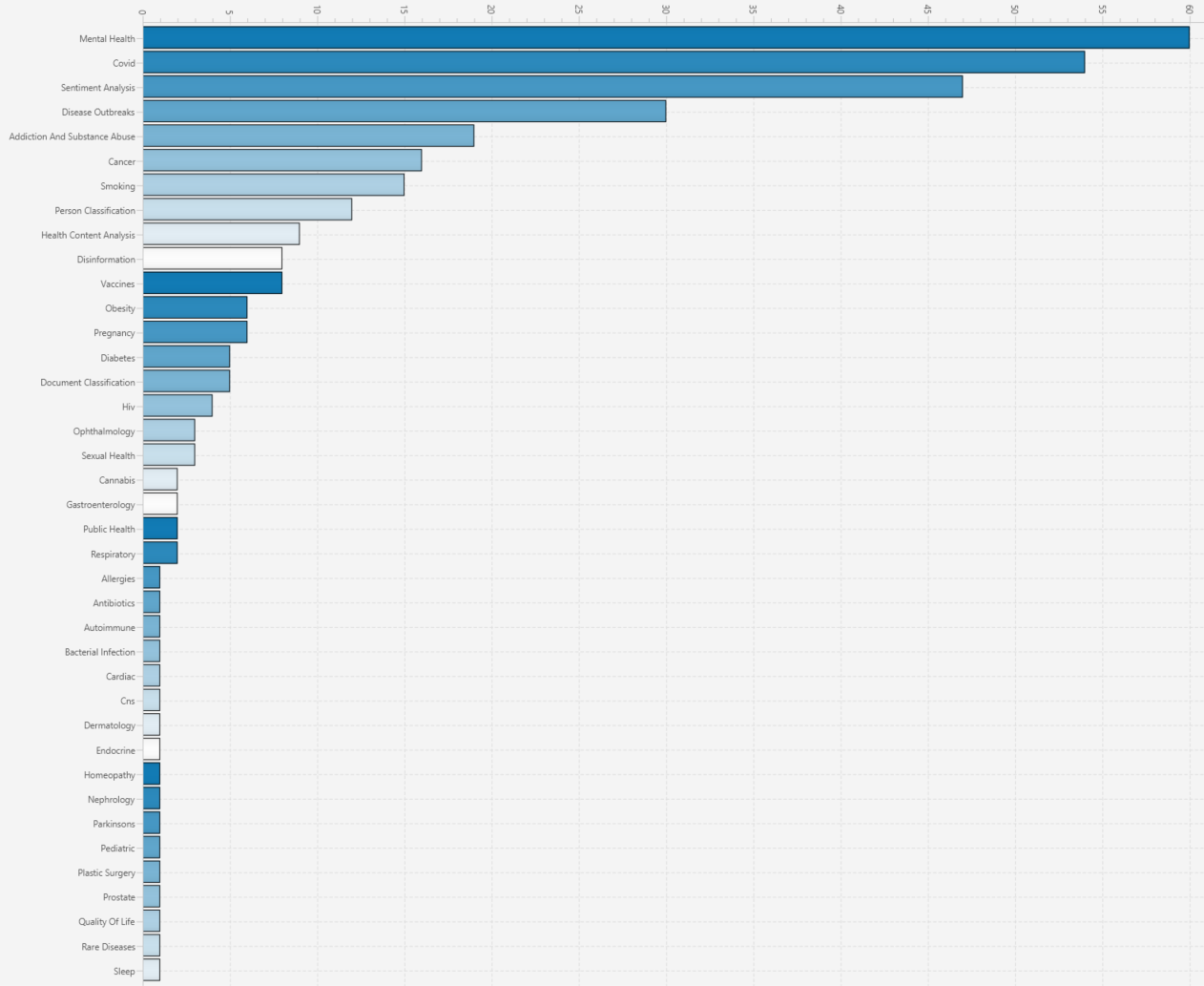

Figure 9: Topic-distribution within the tagged titles and abstracts.

#### Appendix D: Search strategies

##### PubMed

("2005/01/01"[Date - Entry] : "2022/12/31"[Date - Entry])

AND

(("public health"[TIAB] OR "public?health"[TIAB]) AND (forum[TIAB] OR fora[TIAB] OR website[TIAB] OR platform[TIAB] OR data[TIAB] OR resource*[TIAB] OR text*[TIAB])) OR ("clinical trial registration"[TIAB] OR "registry of Clinical Trials"[TIAB] OR "trials registries"[TIAB] OR "trial registries"[TIAB] OR "trials registry"[TIAB] OR "trial registry"[TIAB] OR "trials register"[TIAB] OR Trial Register[MH] OR ClinicalTrials.gov[TIAB] OR ISRCTN[TIAB] OR "IRCT"[TIAB] OR "ChiCTR"[TIAB]) OR

(preprint[TIAB] OR pre-print[TIAB] OR "pre print"[TIAB] OR pre?prints[TIAB] OR preprint[MH] OR medrxiv[TIAB] OR biorxiv[TIAB] OR arxiv[TIAB]) OR ("social media"[TIAB] OR social media[MH] OR twitter[TIAB] OR instagram[TIAB] OR tiktok[TIAB] OR facebook[TIAB] OR LinkedIn[TIAB] OR Snapchat[TIAB] OR Tumblr[TIAB] OR Pinterest[TIAB] OR reddit[TIAB] OR "research gate"[TIAB]) OR ("grey literature"[TIAB] OR "gray literature"[TIAB] OR dissertation*[TIAB] OR ((health[TIAB] OR lifestyle[TIAB]) AND (forum[TIAB] OR fora[TIAB] OR website*[TIAB] OR platform[TIAB])) OR ((government*[TIAB]) AND (website*[TIAB] OR resource*[TIAB])) OR news[TIAB] OR patent*[TIAB]) OR (clinical guideline*[TIAB] OR "health technology assessment"[TIAB] OR hta[TIAB] OR "medical guideline"[TIAB])

AND

((reference* OR literature OR data OR text* OR record* OR article* OR paper* OR post* OR tweet*) AND (retriev* OR search* OR download)) OR ("Automatic extraction"[TIAB] OR "Automated extraction"[TIAB] OR "Automating extraction"[TIAB] OR "data extraction"[TIAB] OR "data-extraction"[TIAB] OR text analys*[TIAB] OR (((text*[TIAB] OR sequence[TIAB]) AND classif*[TIAB]) NOT ((RNA OR nucleotide OR DNA OR protein) AND sequence)) OR datamin*[TIAB] OR data mine*[TIAB] OR textmin*[TIAB] OR "data mining"[MH] OR "literature mining"[MH] OR "text mining"[MH] OR literature mine*[TIAB] OR text mine*[TIAB] OR Data Mining/ OR ((automated[MH] OR automating[MH] OR automation[MH] OR semi-automated[MH] OR semi-automating[MH] OR semi-automation[MH] OR automated[TI] OR automating[TI] OR automation[TI] OR semi?automated[TI] OR semi?automating[TI] OR semi-automation[TI] OR Automation/) AND (pico[TIAB] OR text[TIAB] or literature[TIAB] OR "natural language"[TIAB] OR publication[TIAB] or websit*[TIAB] OR search*[TIAB] or retriev*[TIAB])))

AND

(inference algorithm[TIAB] OR inference algorithms[TIAB] OR statistical relational learning[TIAB] OR statistical relational learning-based approach[TIAB] OR kLog[TIAB] OR "supervised classification"[TIAB] OR "unsupervised classification"[TIAB] OR "un-supervised classification"[TIAB] OR "Information retrieval"[TIAB] OR word embedding[TIAB] OR word2vec[TIAB] OR character embedding[TIAB] OR SCIBERT[TIAB] OR ALBERT[TIAB] OR DistilBERT[TIAB] OR SpanBERT[TIAB] OR RoBERTa[TIAB] OR XLNet[TIAB] OR Transformer-XL[TIAB] OR BIOBERT[TIAB] OR rule based learning[TIAB] OR "rule-base"[TIAB] OR recurrent neural network*[TIAB] OR RNN[TIAB] OR random forest[TIAB] OR radial basis function[TIAB] OR RBFN[TIAB] OR n?ive bayes[TIAB] OR multi-layer perceptron[TIAB] OR perceptron algorithm[TIAB] OR maximum entropy classifier[TIAB] or MaxEnt[TIAB] or Max-Ent[TIAB] OR long short-term memory[TIAB] or LSTM[TIAB] OR bi-LSTM[TIAB] OR latent dirichlet allocation[TIAB] OR topic model[TIAB] OR decision tree?[TIAB] OR conditional random field?[TIAB] OR CRF[TIAB] OR convolutional neural network?[TIAB] OR CNN[TIAB] OR BERT[TIAB] OR Bidirectional Encoder Representations[TIAB] OR "term recognition"[TIAB] or "regular expression"[TIAB] or regex[TIAB] OR support vector machine[TIAB] OR SVM[TIAB] OR ((ontolog*[TIAB] OR "knowledge base"[TIAB] OR knowledgebase[TIAB] or knowledge-base[TIAB]) AND (expert[TIAB] OR database[TIAB])) OR question answering[TIAB] OR reading comprehension[TIAB] OR predictive modelling[TIAB] OR predictive modelling[MH] OR "NLP"[TIAB] OR Natural Language Processing[TIAB] OR Natural Language Processing[MH] OR nlp[TIAB] OR Natural Language Processing/ OR artificial intelligence[TIAB] OR artificial intelligence[MH] OR Artificial Intelligence/ OR neural network[TIAB] OR neural networks[TIAB] OR "active learning"[TIAB] OR "deep learning"[TIAB] OR "supervised learning"[TIAB] OR "semi-supervised learning"[TIAB] OR "transfer learning"[TIAB] OR "unsupervised learning"[TIAB] OR learning algorithm*[TIAB] OR learning algorithm*[MH] OR machine learn*[TIAB] OR machine learn*[MH] OR machine learning/ OR deep learning/ OR supervised machine learning/ OR unsupervised machine learning/ OR neural networks/)

##### Scopus

( PUBYEAR > 2005 ) AND ( ( TITLE-ABS ( "public health" ) OR TITLE-ABS ( public?health ) ) W/1 ( TITLE-ABS ( forum ) OR TITLE-ABS ( fora ) OR TITLE-ABS ( website ) OR TITLE-ABS ( platform ) OR TITLE-ABS ( data ) OR TITLE-ABS ( resource* ) OR TITLE-ABS ( text* ) ) ) OR ( TITLE-ABS ( "clinical trial registration" ) OR TITLE-ABS ( "registry of Clinical Trials" ) OR TITLE-ABS ( "trials registries" ) OR TITLE-ABS ( "trial registries" ) OR TITLE-ABS ( "trials registry" ) OR TITLE-ABS ( "trial registry" ) OR TITLE-ABS ( "trials register" ) OR INDEXTERMS ( "Trial Register" ) OR TITLE-ABS ( clinicaltrials.gov ) OR TITLE-ABS ( isrctn ) OR TITLE-ABS ( irct ) OR TITLE-ABS ( chictr ) ) OR ( TITLE-ABS ( preprint ) OR TITLE-ABS ( pre-print ) OR TITLE-ABS ( "pre print" ) OR TITLE-ABS ( pre?prints ) OR INDEXTERMS ( preprint ) OR TITLE-ABS ( medrxiv ) OR TITLE-ABS ( biorxiv ) OR TITLE-ABS ( arxiv ) ) OR ( TITLE-ABS ( "social media" ) OR INDEXTERMS ( "social media" ) OR TITLE-ABS ( twitter ) OR TITLE-ABS ( instagram ) OR TITLE-ABS ( tiktok ) OR TITLE-ABS ( facebook ) OR TITLE-ABS ( linkedin ) OR TITLE-ABS ( snapchat ) OR TITLE-ABS ( tumblr ) OR TITLE-ABS ( pinterest ) OR TITLE-ABS ( reddit ) OR TITLE-ABS ( "research gate" ) ) OR ( TITLE-ABS ( "grey literature" ) OR TITLE-ABS ( "gray literature" ) OR TITLE-ABS ( dissertation* ) OR ( ( TITLE-ABS ( health ) OR TITLE-ABS ( lifestyle ) ) ( TITLE-ABS ( forum ) OR TITLE-ABS ( fora ) OR TITLE-ABS ( website* ) OR TITLE-ABS ( platform ) ) ) OR ( ( TITLE-ABS ( government* ) ) ( TITLE-ABS ( website* ) OR TITLE-ABS ( resource* ) ) ) OR TITLE-ABS ( news ) OR TITLE-ABS ( patent* ) ) OR ( TITLE-ABS ( "clinical guideline*" ) OR TITLE-ABS ( "health technology assessment" ) OR TITLE-ABS ( hta ) OR TITLE-ABS ( "medical guideline" ) ) AND ( ( reference* OR literature OR data OR text* OR record* OR article* OR paper* OR post* OR tweet* ) W/1 ( retriev* OR search* OR download ) ) OR TITLE-ABS ( "Automatic extraction" ) OR TITLE-ABS ( "Automated extraction" ) OR TITLE-ABS ( "Automating extraction" ) OR TITLE-ABS ( "data extraction" ) OR TITLE-ABS ( data-extraction ) OR TITLE-ABS ( text W/1 analys* ) OR TITLE-ABS ( datamin* ) OR TITLE-ABS ( data W/1 mine* ) OR TITLE-ABS ( textmin* ) OR INDEXTERMS ( "data mining" ) OR INDEXTERMS ( "literature mining" ) OR INDEXTERMS ( "text mining" ) OR TITLE-ABS ( literature W/1 mine* ) OR TITLE-ABS ( text W/1 mine* ) OR INDEXTERMS ( "Data Mining" ) OR ( INDEXTERMS ( automated ) OR INDEXTERMS ( automating ) OR INDEXTERMS ( automation ) OR INDEXTERMS ( semi-automated ) OR INDEXTERMS ( semi-automating ) OR INDEXTERMS ( semi-automation ) OR TITLE ( automated ) OR TITLE ( automating ) OR TITLE ( automation ) OR TITLE ( semi?automated ) OR TITLE ( semi?automating ) OR TITLE ( semi-automation ) OR INDEXTERMS ( automation ) ) AND ( TITLE-ABS ( pico ) OR TITLE-ABS ( text ) OR TITLE-ABS ( literature ) OR TITLE-ABS ( "natural language" ) OR TITLE-ABS ( publication ) OR TITLE-ABS ( websit* ) OR TITLE-ABS ( search* ) OR TITLE-ABS ( retriev* ) ) AND ( TITLE-ABS ( "inference algorithm" ) OR TITLE-ABS ( "inference algorithms" ) OR TITLE-ABS ( "statistical relational learning" ) OR TITLE-ABS ( "statistical relational learning-based approach" ) OR TITLE-ABS ( klog ) OR TITLE-ABS ( "supervised classification" ) OR TITLE-ABS ( "unsupervised classification" ) OR TITLE-ABS ( "un-supervised classification" ) OR TITLE-ABS ( "Information retrieval" ) OR TITLE-ABS ( "word embedding" ) OR TITLE-ABS ( word2vec ) OR TITLE-ABS ( "character embedding" ) OR TITLE-ABS ( scibert ) OR TITLE-ABS ( albert ) OR TITLE-ABS ( distilbert ) OR TITLE-ABS ( spanbert ) OR TITLE-ABS ( roberta ) OR TITLE-ABS ( xlnet ) OR TITLE-ABS ( transformer-xl ) OR TITLE-ABS ( biobert ) OR TITLE-ABS ( "rule based learning" ) OR TITLE-ABS ( rule-base ) OR TITLE-ABS ( recurrent W/1 neural W/1 network* ) OR TITLE-ABS ( rnn ) OR TITLE-ABS ( "random forest" ) OR TITLE-ABS ( "radial basis function" ) OR TITLE-ABS ( rbfn ) OR TITLE-ABS ( "n?ive bayes" ) OR TITLE-ABS ( "multi-layer perceptron" ) OR TITLE-ABS ( "perceptron algorithm" ) OR TITLE-ABS ( "maximum entropy classifier" ) OR TITLE-ABS ( maxent ) OR TITLE-ABS ( max-ent ) OR TITLE-ABS ( "long short-term memory" ) OR TITLE-ABS ( lstm ) OR TITLE-ABS ( bi-lstm ) OR TITLE-ABS ( "latent dirichlet allocation" ) OR TITLE-ABS ( "topic model" ) OR TITLE-ABS ( decision W/1 tree* ) OR TITLE-ABS ( conditional W/1 random W/1 field* ) OR TITLE-ABS ( crf ) OR TITLE-ABS ( convolutional W/1 neural AND network ) OR TITLE-ABS ( cnn ) OR TITLE-ABS ( bert ) OR TITLE-ABS ( "Bidirectional Encoder Representations" ) OR TITLE-ABS ( "term recognition" ) OR TITLE-ABS ( "regular expression" ) OR TITLE-ABS ( regex ) OR TITLE-ABS ( "support vector machine" ) OR TITLE-ABS ( svm ) OR ( ( TITLE-ABS ( ontolog* ) OR TITLE-ABS ( "knowledge base" ) OR TITLE-ABS ( knowledgebase ) OR TITLE-ABS ( knowledge-base ) ) AND ( TITLE-ABS ( expert ) OR TITLE-ABS ( database ) ) ) OR TITLE-ABS ( "question answering" ) OR TITLE-ABS ( "reading comprehension" ) OR TITLE-ABS ( "predictive modelling" ) OR INDEXTERMS ( "predictive modelling" ) OR TITLE-ABS ( nlp ) OR TITLE-ABS ( "Natural Language Processing" ) OR INDEXTERMS ( "Natural Language Processing" ) OR TITLE-ABS ( nlp ) OR INDEXTERMS ( "Natural Language Processing" ) OR TITLE-ABS ( "artificial intelligence" ) OR INDEXTERMS ( "artificial intelligence" ) OR INDEXTERMS ( "Artificial Intelligence" ) OR TITLE-ABS ( "neural network" ) OR TITLE-ABS ( "neural networks" ) OR TITLE-ABS ( "active learning" ) OR TITLE-ABS ( "deep learning" ) OR TITLE-ABS ( "supervised learning" ) OR TITLE-ABS ( "semi-supervised learning" ) OR TITLE-ABS ( "transfer learning" ) OR TITLE-ABS ( "unsupervised learning" ) OR TITLE-ABS ( learning W/1 algorithm ) OR INDEXTERMS ( learning W/1 algorithm* ) OR TITLE-ABS ( machine W/1 learn* ) OR INDEXTERMS ( machine W/1 learn* ) OR INDEXTERMS ( "machine learning" ) OR INDEXTERMS ( "deep learning" ) OR INDEXTERMS ( "supervised machine learning" ) OR INDEXTERMS ( "unsupervised machine learning" ) OR INDEXTERMS ( "neural networks" ) ) AND ( LIMIT-TO ( DOCTYPE , "cp" ) OR LIMIT-TO ( DOCTYPE , "ar" ) ) AND ( LIMIT-TO ( LANGUAGE , "English" ) ) AND ( LIMIT-TO ( SRCTYPE , "p" ) OR LIMIT-TO ( SRCTYPE , "j" ) )

##### Other database searches based on regex: MedRxiv, ACL Anthology, ArXiv, dblp

For the following searches, whole database-dumps were searched on title+abstract level (or title-only for dblp) for each arm. Then, results from each arm were combined with logical ‘AND’ operator to retrieve records mentioning at least one term from each arm.

arm1 <- c("(\\bpublic.health\\b)|(trials?[ -]?regist)|(regist.+of clinical trial)|(clinicaltrials\\.gov)|(isrctn)|(irct)|(chictr)|(pre[- ]?print)|(medrxiv)|(biorxiv)|(arxiv)|(social[ -]?media)|(twitter)|(instagram)|(tik[ -]tok)|(facebook)|(linkedin)|(snapchat)|(tumblr)|(pinterest)|(reddit)|(research[ -]?gate)|(gr[ea]y[ -]?literature)|(dissertation)|(for(um|a))|(website)|(platform)|(lifestyle)|(resource)|(news)|(patent)|(clinical guideline)|(health[ -]?technology[ -]?assessment)|(\\bhta\\b)|(medical guideline)")

arm2 <- c("(automat(ic|ed|ing) extraction)|(data[- ]?extract)|(text[- ]?(analys|min))|(text[- ]?classif)|(data[- ]?(analys|min))|(literature[- ]?min)|(automat(ed|ing|ion).{1,25}(pico|text|literature|natural language|nlp|publication|website|search|retrieve))|((pico|text|literature|natural language|nlp|publication|website|search|retrieve).{1,25}automat(ed|ing|ion))|((text|record|reference|literature|data|article|paper|post|tweet).{1,25}retrieve)|((text|record|reference|literature|data|article|paper|post|tweet).{1,25}search)|((text|record|reference|literature|data|article|paper|post|tweet).{1,25}download)|(search.{1,35}(text|record|reference|literature|data|article|paper|post|tweet))|(retrieve.{1,25}(text|record|reference|literature|data|article|paper|post|tweet))|(download.{1,25}(text|record|reference|literature|data|article|paper|post|tweet))")

arm3 <- c("(inference[- ]?algorithm)|(statistic(al)?[- ]?relational[- ]?learning)|(klog)|(supervised classif)|(information[ -]?retrieval)|(word embedding)|(word2vec)|(character embedding)|((sci|al|bio|distil|span|ro)berta?)|(\bbert\b)|(specter)|(xlnet)|(transformer)|(rule[- ?]base)|(recurrent[- ?]neural[- ?]network)|(rnn|cnn|lstm|mlp|rfbn|max.ent|\\blda\\b|\\bcrf|svm)|(random forest)|(radial basis function)|(n.ive[ -]?bayes)|(layer perceptron)|(perceptron algorithm)|(maximum entropy classif)|(long[ -]?short[ -]?term[ -]?memory)|(latent dirichlet)|(topic[ -]?model)|(decision[ -]?tree)|(conditional[ -]?random[ -]?field)|(convolutional[ -]?neural[ -]?network)|(Bidirectional Encoder Representation)|(term[ -]?recognition)|(regular.expression)|(regex)|(support[ -]?vector[ -]?machine)|(ontolog(y|ies))|(question answering)|(reading comprehension)|(predictive modelling)|(nlp)|(natural language processing)|(artificial intelligence)|(neural[ -]?network)|((active|deep|machine|supervised|transfer)[ -]?learning)|((learn|train).{1,20}algorithm)")

1. <https://www.medhelp.org/> and <https://www.askapatient.com/>
